# Clarifying Values: An Updated and Expanded Systematic Review and Meta-Analysis

**DOI:** 10.1101/2021.01.21.21250270

**Authors:** Holly O. Witteman, Ruth Ndjaboue, Gratianne Vaisson, Selma Chipenda Dansokho, Bob Arnold, John F. P. Bridges, Sandrine Comeau, Angela Fagerlin, Teresa Gavaruzzi, Melina Marcoux, Arwen Pieterse, Michael Pignone, Thierry Provencher, Charles Racine, Dean Regier, Charlotte Rochefort-Brihay, Praveen Thokala, Marieke Weernink, Douglas B. White, Celia E. Wills, Jesse Jansen

## Abstract

**Background:** Patient decision aids should help people make evidence-informed decisions aligned with their values. There is limited guidance about how to achieve such alignment.

**Purpose:** To describe the range of values clarification methods available to patient decision aid developers, synthesize evidence regarding their relative merits, and foster collection of evidence by offering researchers a proposed set of outcomes to report when evaluating the effects of values clarification methods.

**Data Sources:** MEDLINE, EMBASE, PubMed, Web of Science, the Cochrane Library, CINAHL

**Study Selection:** We included articles that described randomized trials of one or more explicit values clarification methods. From 30,648 records screened, we identified 33 articles describing trials of 43 values clarification methods.

**Data Extraction:** Two independent reviewers extracted details about each values clarification method and its evaluation.

**Data Synthesis:** Compared to control conditions or to implicit values clarification methods, explicit values clarification methods decreased the frequency of values-disgruent choices (risk difference -0.04 95% CI [-0.06 to -0.02], p<.001) and decisional regret (standardized mean difference -0.20 95% CI [-0.29 to -0.11], p<0.001). Multicriteria decision analysis led to more values-congruent decisions than other values clarification methods (Chi-squared(2)=9.25, p=.01). There were no differences between different values clarification methods regarding decisional conflict (Chi-squared(2)=6.08, p=.05).

**Limitations:** Some meta-analyses had high heterogeneity. We grouped values clarification methods into broad categories.

**Conclusions:** Current evidence suggests patient decision aids should include an explicit values clarification method. Developers may wish to specifically consider multicriteria decision analysis. Future evaluations of values clarification methods should report their effects on decisional conflict, decisions made, values congruence, and decisional regret.

## Introduction

Shared decision making aims to foster health-related decisions that are both informed by the best available evidence and aligned with what matters to the person or people affected by the decision. [1–4] Individual values are a critical ingredient in high quality individual health decision making. [5–7] What is important to one person might be different from what is important to others, and determining what is important to oneself can be difficult even if one has the appropriate information and evidence at hand. Therefore, patient decision aids should both present evidence appropriately and also support the process of clarifying and expressing patients’ (and, when appropriate, other relevant stakeholders’) values, with the goal of supporting alignment between values and decisions. Within patient decision aids, such support is offered by explicit values clarification methods.

Explicit values clarification methods require users to interact with something such as a worksheet or an interactive website to clarify what matters to them relevant to a health decision. Such methods have been shown to encourage desirable outcomes such as better alignment with patients’ values [8, 9] and reduced decisional regret, the latter particularly among people with lower health literacy. [10] However, explicit values clarification methods are extremely diverse, [11], and there has been little guidance regarding their comparative effects on users’ decision-making processes or outcomes [12], making it difficult for patient decision aid developers to know which explicit method to use. Patient decision aid developers might look towards the preference elicitation literature for guidance, but the guidance available [13] is often tailored towards aggregate level decision making, such as regulatory decisions [14] or health technology assessment [15], not for supporting individual-level decision making.

This updated review sought to build upon previous versions of the International Patient Decision Aids Standards’ Chapter on Values Clarification [16, 17] as well as previous evidence syntheses that have established the advantages of explicit values clarification methods over implicit methods or no values clarification. [8, 9] We sought to advance the science and practice of values clarification methods in three ways. First, we aimed to offer clear definitions and an annotated summary of existing approaches that have been or could be used as values clarification methods. Second, we aimed to synthesize evidence of different techniques’ effects on health decision outcomes. Third, we aimed to foster future evidence by offering researchers a proposed set of outcomes to consider when evaluating the effects of values clarification methods.

### Definitions

Part of the challenge in studying or using values clarification methods is that definitions vary and terms like ‘values’ are used imprecisely in the patient decision support literature. [18, 19] Another challenge is that there is substantial overlap between values clarification methods used in patient decision support and preference elicitation methods used in health economics. To bring clarity to this imprecision and overlap, we adopt working definitions in Table 1 for use in this paper.

**Table 1.** Definitions of Terms

| Term | Definition adopted in this paper |
| --- | --- |
| Values | An umbrella term referring to what matters to an individual relevant to a health decision. Values may be directly relevant to decisions (e.g., "beliefs, feelings, or perceptions regarding attributes of a treatment option") or indirectly relevant (e.g., goals; worldviews; family, religious, or cultural values). [20] Values may be represented qualitatively or, in some cases, quantitatively. This definition is |
|  | deliberately broad. |
| Values clarification | “The process of sorting out what matters to an individual relevant to a given health decision.” [11] This definition emphasizes that what matters to an individual may be broader than attribute-specific values. What matters may also include preferences, concerns, and issues to do with the context of a person’s life within which they would need to implement a decision. |
| Values clarification methods | “Strategies that are intended to help patients evaluate the desirability of options or attributes of options within a specific decision context, in order to identify which option [they] prefer.” [17] |
| Implicit values clarification methods | Strategies for facilitating values clarification that do not require people to interact with anything or anyone; e.g., describing “options in enough detail that clients can imagine what it is like to experience the physical, emotional, and social effects,” [9] or simply encouraging people to think about what matters to them. |
| Explicit values clarification methods | Strategies for facilitating values clarification that require people to interact with something or someone; e.g., filling out a worksheet, using an interactive website, having a semi-structured conversation with another person with the explicit purpose of clarifying values, or engaging in another structured exercise. |
| Preferences | The extent to which a decision option or health state is desirable or acceptable, either in the abstract or in comparison to other options or health states. Preferences may be represented qualitatively or, more commonly, quantitatively. [21] |
| Preference elicitation methods | Processes by which preferences are drawn out. [11] Preference elicitation methods may vary according to the theory informing them. They are highly related to values clarification methods. Although older terms “revealed” and “stated” preference elicitation methods are no longer recommended, readers who encounter these terms in previous preference elicitation literature should note that these may overlap with implicit and explicit values clarification methods, respectively. |

As noted above, we continue to use the term values clarification even though this is sometimes misinterpreted as implying a narrow definition of values. Changing terms makes it difficult for people who are new to a field to connect the dots across decades of previous research. It is clear that previous research in values clarification addressed issues that were broader than valuation of treatment-specific attributes. [16] In this update, we therefore move forward with the older terms, now with more clarity about what they mean in our presentation of the evidence.

### Theoretical Rationale

Our interdisciplinary team determined that the theoretical rationale for values clarification required only a small edit, shown in square brackets, to reflect the focus on explicit methods. Like Fagerlin and colleagues, we assert the theoretical rationale for explicit values clarification methods as being that they, “should aim to [explicitly] facilitate at least one or more of the following six decision-making processes: 1) Identifying options, which can include either the narrowing down of options, or the generation of options that were not offered at the outset, 2) Identifying attributes of the situation and/or the options which ultimately affect the patient’s preference in a specific decision context, 3) Reasoning about options or attributes of options, 4) Integrating attributes of options using either compensatory or both compensatory and non-compensatory decision rules, 5) Making holistic comparisons, and 6) Helping decision makers retrieve relevant values from long-term memory.” [17] Pieterse and colleagues provided theory-based recommendations on processes that values clarification methods could aim to facilitate. [22]

Although reasoning is one of the potential processes supported by values clarification, neither the definition nor the theoretical rationale of values clarification methods requires that people who are being supported in making a personal health decision must rationally deliberate about each option, nor that the goal must always be a fully rational choice. In some decision-making situations, rational deliberation and rational choice may be desired, while in others, they may not. [23, 24]

### Explicit Values Clarification Methods

Table 2 organizes strategies that can be used as explicit values clarification methods in patient decision aids, building upon previously-developed lists of types of values clarification methods [7, 11] and reviews of preference elicitation methods. [25, 26] Methods range from highly structured strategies that can also be used for preference elicitation in the context of health policy decision making to substantially less structured strategies. While not every use of a given method will be exactly the same, we deemed them functionally similar in terms of how they might be used and what the user experience might be in a patient decision aid. Patient decision aids may use multiple strategies. For example, a user may be asked to use a rating scale or visual analog scale whose values are then used in a decision analytic model.

**Table 2.** Values Clarification Methods

| Method | Description |
| --- | --- |
| Adaptive Conjoint Analysis (example [27]) | The user rates a series of sets of attributes and their levels, where choices presented are tailored to earlier answers. |
| Allocation of Points (example [28]) | The user has a “budget” to “spend” on decision attributes, according to their importance. |
| Analytical Hierarchy Process (example [29]) | The user is asked to compare sets of options relative to predefined decision criteria. |
| Best-Worst Scaling (example [30]) | The user is asked to indicate the best and the worst in sets of options with different attributes and levels. |
| Decision Analysis or Multicriteria Decision Analysis (umbrella term*) (resource [31, 32]) | The user is asked to directly indicate the extent to which a decision attribute or outcome matters to them or how good or bad they deem it to be. These values are then used in a model that calculates alignment between what matters to the user and the available decision options. |
| Discrete Choice Experiments (example [33]) | The user is asked to make a series of choices between two (or more) alternatives, where each alternative is characterized by attributes and their associated levels. |
| Open Discussion (example [34]) | The user makes a list and/or discusses what matters to them in an unstructured or semi-structured discussion. |
| Pros and Cons (resource [35]) | The user lists advantages (pros) and disadvantages (cons) of options and/or indicates the relevance (‘this matters to me’) or importance (e.g., on a Likert scale) of each advantage or disadvantage |
| Ranking (example [36]) | The user is asked to place attributes in order of importance, relative to each other. |
| Rating Scales (example [37]) | The user indicates the importance of an attribute on a visual analog scale (e.g., paper-based visual analog scale, online slider) or Likert scale approximating a visual analog scale. If the rating is then used to calculate and show which option fits best, the method is classified as (multicriteria) decision analysis. |
| Social Matching (example [38]) | The user “observes different characters’ decisions and/or decision-making processes and identifies 1 or more characters” with whom they identify. [11] |
| Standard Gamble (example [39]) | The user indicates their willingness to accept a certain risk of death in order to avoid a particular health state by choosing between the certainty of living in that health state for the remainder of their life versus a gamble between two possible outcomes: life in a state of optimal health, with probability p, or immediate death, with probability (1–p). |
| Time Tradeoff (example [39]) | The user indicates how many years in their current health state they would be willing to ‘trade off’, in order to regain full health. |

\*Multicriteria decision analysis or decision analysis is an umbrella term. It encompasses some of the other, more specific categories (e.g., discrete choice experiments, best-worst scaling.) When applicable, we use the more specific, narrower categories. Otherwise, we use the umbrella term “multicriteria decision analysis,” or, for brevity in figures, “decision analysis.” Additionally, although within multicriteria decision analysis, the user may be asked to rate attributes on rating scales, what distinguishes multicriteria decision analysis from other methods such as rating scales is that the model calculates how well or poorly the options align with what matters to a user.

## Methods

Our overall methods were guided by the Cochrane Handbook. We report according to the Preferred Reporting Items for Systematic Reviews and Meta-Analyses (PRISMA) [40] guidelines.

### Eligibility criteria

We included published reports of comparative evaluations of explicit values clarification methods, whether they were called ‘values clarification methods’ in the publications or not. This meant that we included trials of preference elicitation methods that had been trialed as values clarification methods; for example, multi-criteria decision analysis or discrete choice experiments. We included evaluations using comparative methods; i.e., randomized controlled trials or randomized experiments of one or more values clarification methods. The comparisons could be one or more values clarification methods compared to a control method, or compared to each other. Because we sought to understand the effects of values clarification methods, we excluded evaluations using descriptive study designs (e.g., acceptability and feasibility study, development study), observational study designs (e.g., reporting outcomes before and after use of a values clarification method), and reports of values clarification methods that did not evaluate the method independently of the patient decision aid in which it was used. Randomized experiments comparing one or more values clarification methods had to use distinctly different methods, meaning that more than the content or presentation of information in the values clarification method varied.

We did not apply language restrictions. We applied date restrictions to the portion of the review for which we had already conducted a systematic review (i.e., evaluations of values clarification methods that used the term ‘values clarification’. [12, 17] Specifically, for this subgroup, we added articles indexed or published starting in 2014 to the existing set of articles indexed or published prior to 2014 that we had already identified using the same search strategy. We applied no date restrictions to the new, expanded portion of the review (i.e., evaluations of values clarification methods that did not use the term ‘values clarification’).

### Information sources

We performed a systematic literature search in MEDLINE, EMBASE, Web of Science, the Cochrane Library, and CINAHL.

### Search strategy

We developed a draft search strategy in collaboration with an information specialist (FB, see Acknowledgments). Search strategies for each database are shown in Online Appendix 1. We reviewed search strategies with all authors to ensure they were inclusive of relevant preference elicitation methods that might be used for values clarification. We conducted hand searches by reviewing articles that cited the previous version of these standards (values clarification chapter) or a previous systematic review of values clarification methods.

### Study records: Data management

We managed data with Covidence (covidence.org, Melbourne, Australia), reviewing data records at regular team meetings.

### Study records: Selection process

Two independent reviewers (SC, MM, TP, CR, CRB) screened titles and abstracts to assess potential relevance, with a third reviewer adjudicating discrepancies and discussions of questions and points of disagreement at regular team meetings. Two independent reviewers then reviewed the full text of all articles deemed potentially relevant based on their title and abstract. Discrepancies in inclusion and exclusion at full text were adjudicated through team discussions at regular meetings until we reached consensus.

### Study records: Data collection process

Two independent, trained research team members (SC, MM, TP, CR, CRB) extracted data from each article using a standardized and pilot-tested data extraction form based on a previous form [12] and adapted to this review. We resolved lack of agreement through discussion until consensus was reached. We contacted authors to collect any needed data that they did not report or were unable to report in their publication.

### Data items

Regarding study participants, we recorded the sample size for control and intervention groups along with basic inclusion and exclusion criteria and whether or not they were making the actual decision or if the study was hypothetical. We defined a hypothetical scenario as one in which people are asked (explicitly or implicitly) to imagine that they are in a certain situation or facing a certain decision. We defined a real scenario as one in which people are facing a decision (e.g., because they have received a diagnosis) or are members of a population likely to face the decision in the near term (e.g., parents of children eligible to receive vaccines within the coming months.)

Regarding interventions, we recorded the type of explicit values clarification method as listed in Table 2. We also recorded specific characteristics of each values clarification method, namely, whether it explicitly requires the user to engage with tradeoffs (i.e., considering which potential harms are acceptable in exchange for their associated potential benefits), whether it explicitly shows the user the correspondence between their options and what they value, and which, if any, theoretical or conceptual framework underpins it. Where relevant, we recorded whether a variable was collected via self-report, meaning whether responses were completed by participants themselves, or by independent researchers based on direct observation, including coded qualitative data.

For comparators (controls) we recorded whether the comparator was no values clarification method or an implicit method, and treated both as equivalent controls. The Cochrane review of patient decision aids specifies that all patient decision aids must contain implicit values clarification methods at minimum [9] and it is accordingly rare to have patient decision aids that do not present potential benefits and harms of options in organized ways. In other words, in the context of patient decision aids, there is no meaningful distinction between implicit methods and no values clarification. The different terminology is simply a function of how authors choose to name their control. We also recorded studies that compared different types of explicit values clarification methods to each other.

### Outcomes

Whenever such data were available, we extracted data regarding values congruence as our primary outcome, as well as secondary outcomes: decision readiness (worry, decision uncertainty, decision-making preparation, knowledge); decisional conflict; decision made; post-decision and post implementation health and well-being (decisional regret, longer-term health outcomes). Following data extraction by pairs of trained reviewers (SC, MM, TP, CR, CRB), three authors (HOW, SCD, JJ) mapped all outcomes into broad outcome groups: worry (including perceived risk), decision uncertainty (not including decisional conflict), decisional conflict (decisional conflict scale or any subscales), decision-making preparation (including self-efficacy for decision-making), beliefs (including beliefs about the condition or underlying decision structure), knowledge, values (including reported utilities), shared decision making, effects on communication (including quality, length, or existence of communication), satisfaction with care, preferences, decision (choice made and implemented) or decisional intent (choice intended, or made and not yet implemented), values congruence, informed decision making, post-decision feelings (including satisfaction, regret), post-decision health, and user assessment of the intervention (including acceptability, satisfaction, perceived balance.) We conducted meta-analyses on primary outcome values congruence and secondary outcome decisional conflict, as these outcomes had sufficient studies to do so.

### Risk of bias in individual studies

Independent, trained research team members assessed risk of bias for each study using methods as defined in the Cochrane Handbook, section 8.5. [41] We conducted quantitative data syntheses with and without studies identified as being at high risk of bias to determine the sensitivity of overall findings to these studies.

### Data synthesis

We synthesized frequency-based results (e.g., how many values clarification methods reflect a given design) descriptively. To synthesize effects on outcomes, we pooled all experiments that evaluated a values clarification method against no values clarification method or an implicit method. For multi-armed studies in which the comparison of a decision aid with and without a values clarification method included an arm that was not relevant to our comparison of interest (for example, an information booklet serving as a control condition in an evaluation of the decision aid) we ignored the third arm. For multi-armed studies containing two or more different values clarification methods and one arm of implicit values clarification or control, we considered each comparison of a values clarification method against implicit values clarification, meaning that each of the multi-armed studies included in this review contributed multiple comparisons to the pooled set.

To meta-analyze results for values congruence, we pooled results using risk differences and applying a random effects model. We extracted dichotomous data indicating the frequency (i.e., number of events and sample size) of values discongruent decisions. To meta-analyze results for decisional conflict, we pooled results using standardized mean differences applying a random effects model. We extracted data on total scores on the Decisional Conflict Scale. We explored and reported consistency using Higgins I^2 [42]. We used the Cochrane Risk of Bias tool to assess study bias along 7 domains as well as to assess an overall risk of bias. Where data permitted, we conducted subgroup meta-analyses of different types of explicit values clarification methods and of explicit values clarification methods that do and do not contain specific design features already identified in previous work [11], namely, whether the method explicitly requires the user to engage with tradeoffs, whether it explicitly provides the user with the implications of what they value, and which, if any, theoretical or conceptual framework underpins it. We used p=0.05 as a threshold for statistical significance and conducted analyses in RevMan, version 5.4.

## Results

### Articles identified

Out of 30,648 records screened at the title and abstract stage and 279 screened at the full text stage, we identified 33 articles that met our inclusion criteria describing trials of 43 values clarification methods. Twenty-four of the articles were new articles identified in this update of IPDAS. We excluded 2 of the articles previously included in the IPDAS Values Clarification Chapter because they did not meet our revised inclusion criteria requiring randomized controlled trials and instead reported, for example, pre-post study designs. The PRISMA diagram of included articles is shown in Figure 1.

**Figure 1.**
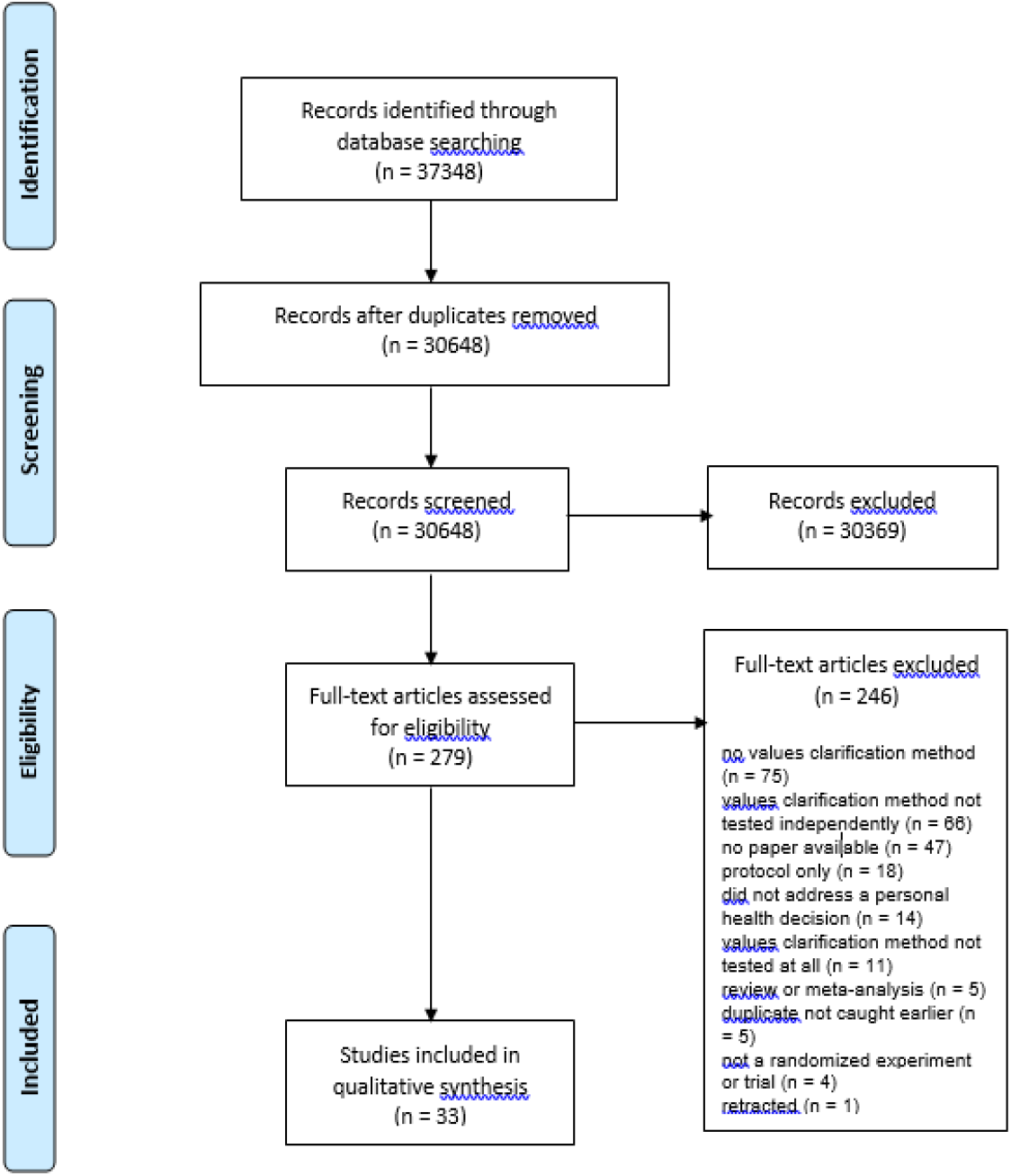
PRISMA Diagram

The decision context varied across studies. Out of the 43 included trials, 25 (58%) addressed treatment decisions, 9 (21%) screening decisions, 4 (9%) prevention, 3 (7%) genetic testing, and 2 (5%) diagnostic testing. Thirteen of the 43 trials (30%) centered around a yes/no decision to take an option or not, 18 (42%) a choice between two or more options and 12 (28%) both a yes/no and a choice between two or more options. Most decisions (22/43, 51%) were real decisions, meaning that the person was making this decision in their actual life. The rest were hypothetical (18/43, 42%) or it was not entirely clear whether the decision was real or hypothetical (3/43, 3%). The most commonly-reported outcomes were decisional conflict and/or its subscales (29/43, 67%), decision and/or decisional intentions (22/43, 51%), knowledge (13/43, 30%), and values congruence (12/43, 28%).

As shown in the overview of included studies in Table 3, there was substantial diversity in the types of values clarification methods used. Decision analysis or multicriteria decision analysis was the most commonly trialed method. Full study details are available in Online Appendix 2.

**Table 3.** Study Details

| Type(s) of Values Clarification Method(s) | Study | Population* | Decision | Summary of findings |
| --- | --- | --- | --- | --- |
| Adaptive Conjoint Analysis | de Achaval and others 2012 [43] | n=208 people with knee osteoarthritis | Whether to receive medication and therapy or total knee arthroplasty | Values clarification method decreased decisional conflict and required more intense cognitive involvement. |
| Adaptive Conjoint Analysis | Fraenkel and others 2007 [44] | n=87, age at least 60 years old, self report of pain involving one or both knees on most days of the month | Choice between five treatments for knee pain | Values clarification method increased self-confidence in and preparation for shared decision-making, and increased arthritis self-efficacy. |
| Adaptive Conjoint Analysis | Hess and others 2015 [45] | n=374 women aged 18 years or older with abnormal uterine bleeding & potential candidates for either surgical or medical treatment | Whether or not to be treated for abnormal uterine bleeding, and, if yes, which treatment to undertake | Values clarification method did not reduce decision regret nor improve treatment satisfaction. |
| Adaptive Conjoint Analysis | Hutyra and others 2019 [46] | n=200 people between 18 and 35 years of age at risk for experiencing a first-time anterior shoulder dislocation | Operative or nonoperative treatment for first-time anterior shoulder dislocation | Values clarification method increased alignment between patients' treatment decisions and evidence-based recommendations. |
| Adaptive Conjoint Analysis | Jayadevappa and others 2015 [47] | n=743 people with newly diagnosed localized prostate cancer | Choice between six options for early stage prostate cancer | Values clarification method improved satisfaction with care, satisfaction with decision, reduced regrets, and aligned treatment choice with risk category. |
| Allocation of Points | Witteman and others 2020 [48] | n=817 adults asked to imagine they had been diagnosed with colon cancer | Choice between two hypothetical surgeries for colon cancer | Values clarification method (strategy 6b in paper) increased values congruence and reduced decisional conflict. |
| Analytical Hierarchy Process | Myers and others 2003 [49] | n=199 men aged 50 to 69 with no personal history of prostate cancer/benign prostate hyperplasia | Whether or not to be screened for prostate cancer | Values clarification method decreased rates of prostate cancer screening. Race/ethnicity analyses showed African American men increased screening while white men decreased screening. |
| Analytical Hierarchy Process | Myers and others 2005 [50] | n=242 African-American men, 40-69 years of age and no history of prostate cancer | Whether or not to be screened for prostate cancer, and if yes, choice of method/extent of screening | Values clarification method increased prostate cancer screening. |
| Best-Worst Scaling | Shirk and others 2017 [51] | n=122 men with incident localized prostate cancer | Choice between three options for incident localized prostate cancer | Values clarification method decreased decisional conflict. |
| Decision Analysis** | Bekker and others 2004 [52] | n=106 pregnant women receiving a screen positive maternal serum screening result | Whether or not to have a prenatal diagnosis for Down syndrome | Values clarification method helped women make more informed prenatal diagnosis decisions. |
| Decision Analysis** | Clancy and others 1988 [53] | n=1280 resident and faculty physicians unvaccinated against Hepatitis B | Choice between three options to manage risk of hepatitis B | Values clarification method resulted in greater action-taking (screening or vaccination.) |
| Decision Analysis** | Feldman-Stewart and others 2012 [54] | n=156 people with newly diagnosed prostate cancer | Choice between more than five main options for early stage prostate cancer | Values clarification method increased preparation for decision making and decreased decision regret. Decision conflict decreased with and without values clarification method. |
| Decision Analysis** | Hopkin and others 2019 [55] | n=349 adults asked to imagine that they had to choose a statin | Choice between five commonly-used statins | Values clarification method reduced decisional conflict and increased levels of preparation for decision making. |
| Decision Analysis** | Montgomery and others 2003 [56] | n=217 adults aged 30-80 years with newly diagnosed hypertension | Whether or not to start drug therapy for hypertension | Values clarification method increased knowledge and reduced total decisional conflict by significantly reducing scores on uninformed, unclear values and unsupported subscales and somewhat reducing scores on uncertainty subscale. Values clarification method did not influence scores on decision quality subscale, nor did it change state anxiety, decision intention, nor ultimate decision. |
| Decision Analysis** | Montgomery and others 2007 [57] | n=742 pregnant women with one previous lower segment caesarean section | Choice of planned mode of delivery | Values clarification method reduced decisional conflict and increased frequency of having a vaginal birth. |
| Decision Analysis** | Witteman and others 2015 [58] | n=407 parents who make medical decisions for at least one child aged 6 months to 18 years and whose child had not yet received the flu vaccine | Whether their child would receive a vaccine against influenza this flu season | Values clarification method had no effect on values congruence. Values clarification method combined with best practices in risk communication increased intentions to vaccinate, particularly among participants who had not had their children vaccinated against influenza in the past 5 years. |
| Decision Analysis** | Witteman and others 2020*** [48] | n=1731 adults asked to imagine they had been diagnosed with colon cancer deciding between two treatment options | Choice between two hypothetical surgeries for colon cancer | Values clarification method (strategies 2a, 2a+2b, 6c, 6b+6c in paper) increased values congruence and reduced decisional conflict this was when measured (strategies 6c, 6b + 6c in paper). |
| Discrete Choice Experiment | Brenner and others 2014 [59] | n=615 people between the ages of 50 and 75 at average | Whether or not to be screened for colorectal cancer, and, if yes, | Values clarification method influenced choice of most important screening test attribute but did not affect unlabeled test |
|  |  | risk for colorectal cancer | which screening test to use | preference, values clarity, nor intent to be screened. |
| Discrete Choice Experiment | Pignone and others 2012 [60] | n=104 adults 48-75 at average risk for colon cancer | Whether or not to be screened for colorectal cancer, and, if yes, which screening test to use | Values clarification method influenced choice of most important attribute, but did not affect values clarity, intent to be screened, or choice of unlabeled screening test. |
| Discrete Choice Experiment | Pignone and others 2013 [61] | n=604 men aged 50-70 at average risk of prostate cancer | Whether or not to be screened for prostate cancer | Values clarification method slightly reduced choice of dying as the most important attribute and increased unlabeled PSA-like screening option but did not influence intent to be screened. |
| Open Discussion | Au and others 2012 [62] | n=306 people with chronic obstructive pulmonary disease | Preferences for end-of-life care | Values clarification method helped identify what mattered to patients regarding end-of-life care and communication. Quality of communication improved. |
| Open Discussion | Epstein and others 2018 [63] | n=99 people with advanced gastrointestinal cancer | Choice between options for end-of-life care | Values clarification method improved communication about future medical cancer care but had no effect on decisional conflict or well-being and increased distress. |
| Open Discussion | Kennedy and others 2002 [64] | n=894 women with uncomplicated menorrhagia | Choice between treatment options for menorrhagia | Values clarification method resulted in minimal improvements in self-reported health status, lower use of a more invasive treatment, higher patient satisfaction, more frequent clinician perceptions of "longer than usual" consultations, and lower overall costs. Providing information alone did not affect treatment choices. |
| Open Discussion | Lerman and others 1997 [65] | n=700, Women aged 18-75 years who had had at least one first-degree relative with breast and/or ovarian cancer | Whether or not to provide a blood sample for BRCA1 testing in the future | Values clarification method increased the perceived importance of the limitations and risk of BRCA1 testing and decreased the perceived importance of the benefits of BRCA1 testing. No effect of values clarification method on intent to treat. |
| Open Discussion | Matheis-Kraft and others 1997 [66] | n=60 women over age 70 with at least one family member or friend who might act as their proxy to make decisions about life-sustaining treatment | Preferences for care in case of decisional incapacity | Values clarification method's effectiveness or lack thereof depended on which statistic (kappa or percent agreement) was used to measure concordance between women and proxies. |
| Pros and Cons | Abhyankar and others 2010 [67] | n=30 healthy women asked to imagine having been diagnosed with breast cancer, undergone lumpectomy and suggested chemotherapy by their doctor | Choice between having standard adjuvant chemotherapy or taking part in a clinical trial testing a new chemotherapy for early stage breast cancer | Values clarification method resulted in more use of personal values when evaluating attributes of options, somewhat less ambivalence, less uncertainty and did not change preferred option. |
| Pros and Cons | O'Connor and others 1999 [68] | n=201 women aged 50-69 who had never used hormone therapy | Whether or not to take hormone replacement therapy after menopause | Values clarification method had no effect on clarity of values, values congruence, total decisional conflict, other subscales of Decisional Conflict Scale, nor acceptability of intervention. |
| Pros and Cons | Paquin and others 2018 [69] | n=1000 people aged 18-44 who were pregnant or whose | Whether or not to use genomic sequencing to identify genetic | Values clarification method decreased parental beliefs against genomic sequencing. |
|  |  | partner was pregnant or planning to become pregnant in the next 2 years | variants in one's child |  |
| Pros and Cons | Peinado and others 2020 [10] | n=1000 people aged 18-44 who were pregnant or whose partner was pregnant or planning to become pregnant in the next 2 years | Whether or not to enroll their newborn child in a medical research study that would involve screening for genetic conditions | Values clarification method decreased decisional regret and increased clarity of personal values but had no effect on overall decisional conflict nor on intent to have one's child tested. |
| Pros and Cons | Witteman and others 2020 [48] | n=772 adults asked to imagine they had been diagnosed with colon cancer | Choice between two hypothetical surgeries for colon cancer | Values clarification method (strategy 4b in paper) reduced decisional conflict but did not change values congruence. |
| Rating Scales | Garvelink and others 2014*** [70] | n=271 healthy women | Whether or not to undergo fertility preserving procedures prior to cancer treatment | Values clarification method had no effect on knowledge nor decisional conflict. |
| Rating Scales | Kuppermann and others 2014 [71] | n=710 pregnant women who had not yet undergone screening or diagnostic testing for fetal aneuploidy in the current pregnancy | Whether or not to have any screening or diagnostic testing for fetal aneuploidy; if screening or testing is desired, whether to start with screening or with invasive diagnostic testing; and which specific screening and/or diagnostic test(s) to undergo. | Values clarification method increased patient knowledge and resulted in less invasive prenatal test use and more informed choices. Values clarification method did not change decisional conflict nor decisional regret. |
| Rating Scales (with and without decision analytic summary) | Feldman-Stewart and others 2006 [72] | n=90 male volunteers asked to imagine that they had just been diagnosed with early-stage prostate cancer | Choice between four options for early stage prostate cancer | Participants preferred values clarification method with decision analytic summary over values clarification method without summary and no values clarification method. |
| Rating Scales | Witteman and others 2020 [48] | n=785 adults asked to imagine they had been diagnosed with colon cancer | Choice between two hypothetical surgeries for colon cancer | Values clarification method (strategy 6a in paper) reduced decisional conflict but did not change values congruence. |
| Rating Scales + Ranking | Brenner and others 2014 [59] | n=614 people between the ages of 50 and 75 at average risk for colorectal cancer | Whether or not to be screened for colorectal cancer, and, if yes, what screening test to use | Values clarification method increased the importance placed on risk reduction as an important attribute but did not affect unlabeled test preference, values clarity, nor intent to be screened. |
| Rating Scales + Ranking | Pignone and others 2012 [60] | n=104 adults 48-75 at average risk for colon cancer | Whether or not to be screened for colorectal cancer, and, if yes, what screening test to use | Values clarification method influenced choice of most important attribute, but did not affect values clarity, intent to be screened, or choice of screening test. |
| Rating Scales + Ranking | Pignone and others 2013 [61] | n=609 men aged 50-70 at average risk of prostate cancer | Whether or not to be screened for prostate cancer | Values clarification method increased the importance placed on dying but did not influence intent to be screened. |
| Rating Scales + Ranking | Sheridan and others 2010 [73] | n=137 men aged 45-80 with no prior history of cardiovascular | Whether or not to initiate behaviours to prevent coronary heart disease, and, if so, | Values clarification method had no effect. Decisional conflict, perceived values congruence, and self-efficacy for health behaviours improved with and without |
|  |  | disease | which behaviours | values clarification. Behavioural intentions did not change. |
| Time Tradeoff + Rating Scales | Frosch and others 2008 [74] | n=611 men older than 50 years | Whether or not to be screened for prostate cancer | Values clarification method increased cancer knowledge scores and decreased decisional conflict. |

\*n is given for the study as a whole. See supplementary appendix for further details about each study.
\*\*Decision analysis or multicriteria decision analysis is an umbrella term. It encompasses some of the other, more specific categories (e.g., discrete choice experiments, best-worst scaling.) Throughout the paper, when applicable, we use the more specific, narrower categories. Otherwise, we use the umbrella term “multicriteria decision analysis,” or, for brevity in figures, “decision analysis.”
\*\*\*Garvelink and colleagues 2014 and Witteman and colleagues 2020 each reported multiple experiments testing values clarification methods that did not differ in type nor in outcomes. Pooled results are therefore presented here.

### Quality assessment

Overall study quality was acceptable, with the majority of studies at low risk of bias on most elements. Eight studies were deemed to be at high risk of bias on one element; the majority in Blinding of Participants and Personnel (Performance Bias). Eighteen additional studies were deemed unclear on this element. Blinding of Outcome Assessment (Detection Bias) was the next most common source of potential bias, with 1 study at high risk of bias and 20 more unclear. Full details of risk of bias assessments are available in Online Appendix 3.

### Values Congruence

As shown in Figure 2a, included explicit values clarification methods, as a group, increased values congruence. Eleven out of 43 trials (26%) reported the number of people who made values-congruent or values-disgruent decisions. The pooled risk difference of making a values-disgruent decision when using one of the trialed values clarification methods was -0.04 95% CI [-0.06 to -0.02], p<.001. The I^2^ of 28% indicates a relatively low level of statistical heterogeneity.

**Figure 2a.**
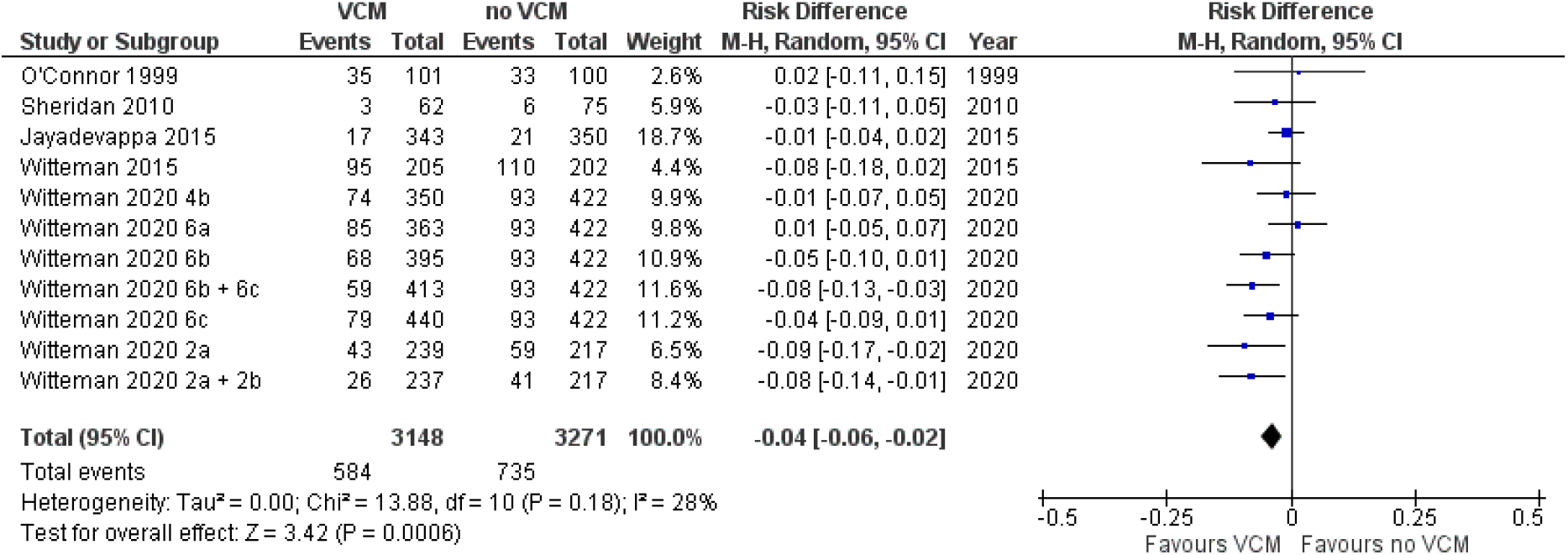
Values Congruence*: Overall (All Values Clarification Methods Together) *Events refer to values-disgruent decisions. The meta-analysis synthesizes the risk across trials of making a values-disgruent decision.

**Figure 2b.**
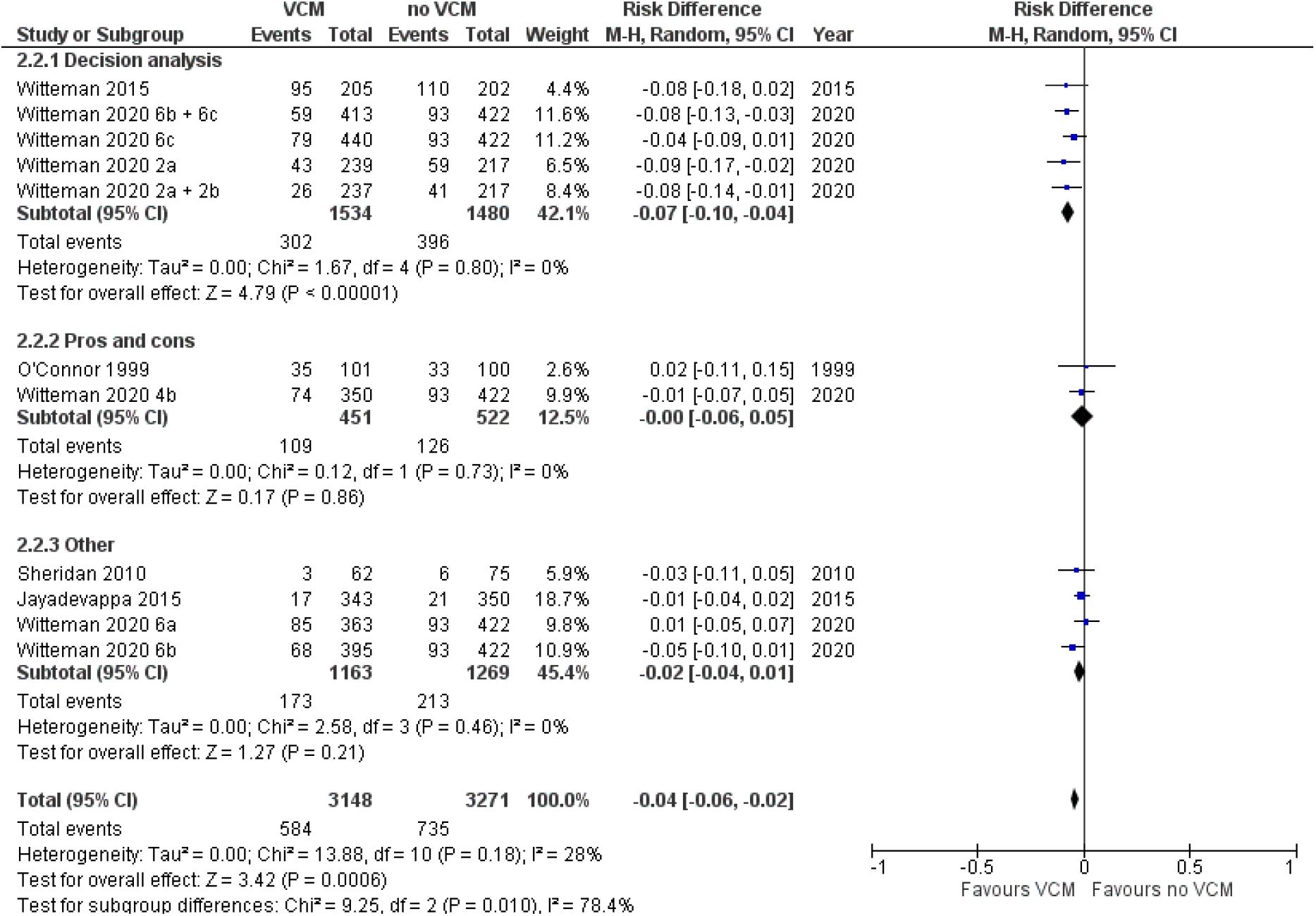
Values Congruence* by Type of Values Clarification Method *Events refer to values-disgruent decisions. The meta-analysis synthesizes the risk across trials of making a values-disgruent decision.

Figure 2b shows a statistically significant subgroup difference by type of values clarification method. The results suggest that decision analysis is more likely to encourage values-congruent decisions compared to other explicit values clarification methods within this set of trials (Chi-squared(2)=9.25, p=.01). The results show no significant subgroup differences by trade-offs, implementation, theory or by implication of the decision. (See Online Appendix 3.) There were no studies in this analysis with a high risk of bias.

### Decisional Conflict

As shown in Figure 3a, explicit values clarification methods decrease decisional conflict. For the 14/43 (33%) trials for which we had complete data, the pooled standardized mean difference for decisional conflict was - 0.20 95% CI [-0.29 to -0.11], p<0.001. The I^2^ of 67% represents moderate to high statistical heterogeneity. Figure 3b shows there was no significant subgroup difference by type of values clarification method (Chi-squared(2)=6.08, p=.05). We found no significant subgroup differences by trade-offs, implementation, theory, implication, nor risk of bias (see Online Appendix 3).

**Figure 3a.**
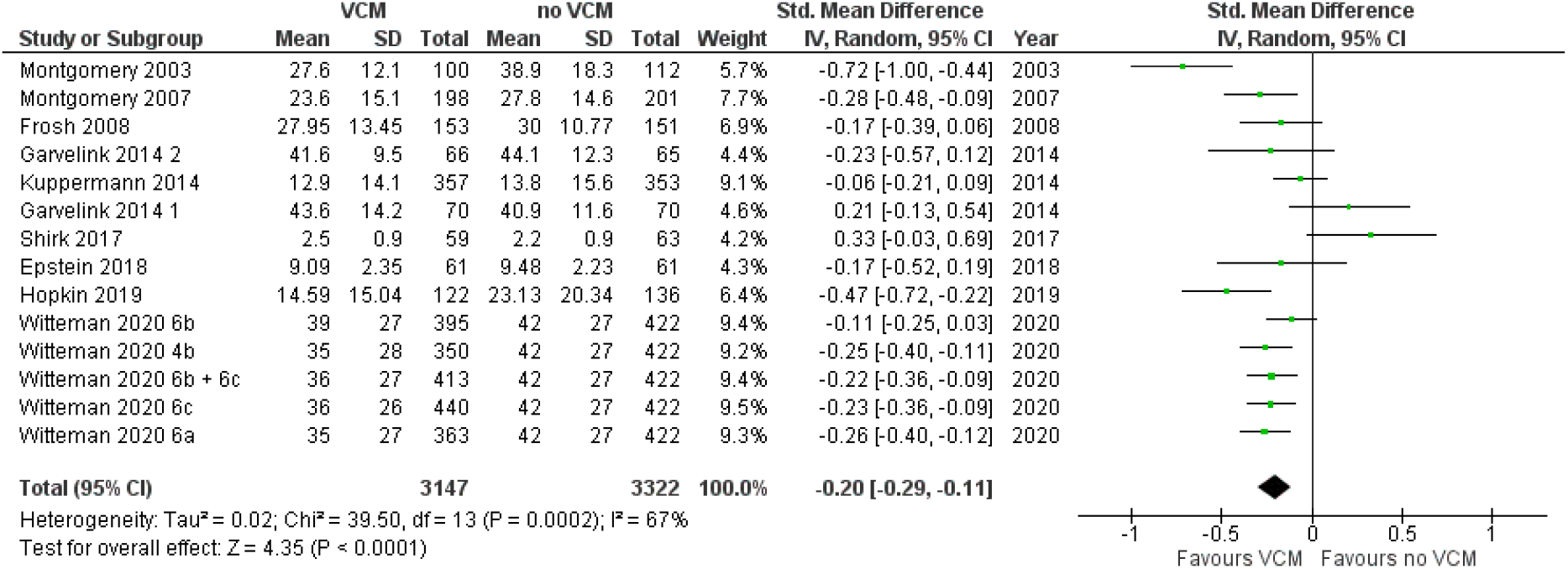
Decisional Conflict: Overall Measure

**Figure 3b.**
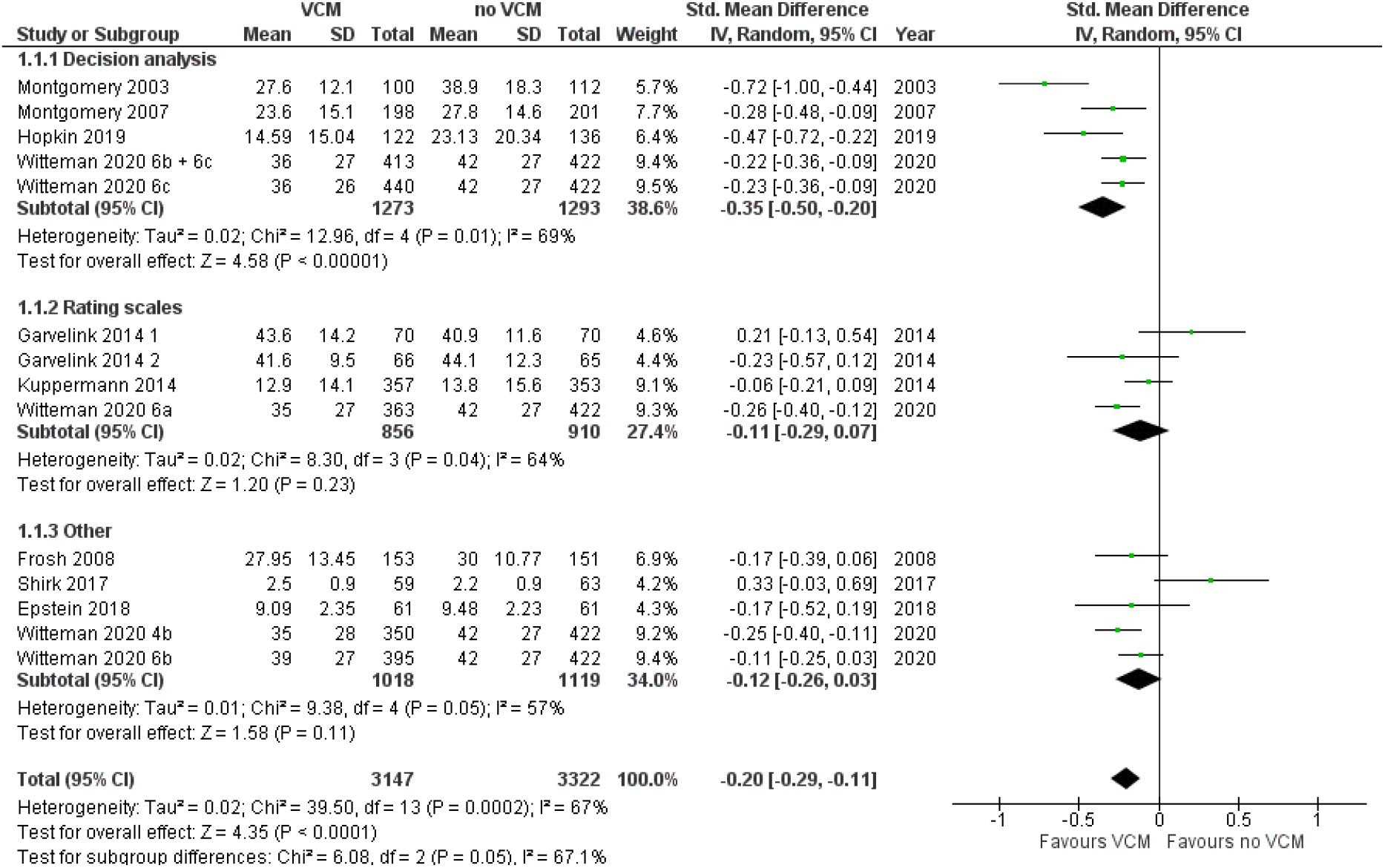
Decisional Conflict by Type of Values Clarification Method

### Head-to-Head Evaluations of Values Clarification Methods

The five studies that compared values clarification methods to each other reported findings that align with the findings of our meta-analyses. Methods that provided users with explicit feedback regarding how the decision options align with their stated values led to somewhat better outcomes, including greater values congruence. [48] When asked to compare methods to each other, study participants also preferred a values clarification method that explicitly showed them how the decision options align with their stated values. [72] Different values clarification methods yielded different patterns of attribute importance. [59–61] Brief summaries of each study are available in Online Appendix 3.

## Discussion

Overall, our systematic review and meta-analyses confirm that explicit values clarification methods improve decision outcomes, notably by increasing values congruence and decreasing decisional conflict. Patient decision aids should include an explicit values clarification method.

While the best explicit values clarification method may depend on context—for example, urgent versus routine care or the extent to which a decision has a clear set of decision attributes—our analyses suggest that patient decision aid developers may wish to consider methods that draw on multicriteria decision analysis. The apparent advantages of such methods shown in our analyses may reflect similarities between the process and the outcome. In other words, increased values congruence yielded by decision analytic methods may be a function of the ways in which such methods transparently show people how their options align with their stated values. We additionally caution that when these methods use pre-specified attributes, there might not be the flexibility for users to add new attributes, highlighting the importance of research to inform attribute selection. We acknowledge that some researchers have argued that health professionals having an unhurried, high-quality conversation with patients may be a preferred approach for at least some patients, especially when decision attributes are many and varied. However, in this systematic review, trials of Open Discussion values clarification methods did not demonstrate strong results, suggesting that such an ideal may be difficult to achieve.

To advance further knowledge on the merits and pitfalls of different values clarification methods, we recommend that authors of future trials of values clarification methods report four outcomes: decisional conflict, decision or decision intention, values congruence, and decisional regret. When possible, authors should make use of validated scales that have good psychometric properties and are commonly reported, as this facilitates evidence synthesis.

Decisional conflict should be assessed before people make the decision, using a version of the Decisional Conflict Scale. [75, 76] Decisions or decision intentions should be assessed when the decision is made.

Values congruence should be assessed once the decision is made. We acknowledge that including values congruence as an outcome brings both measurement and conceptual issues. Measurement issues exist because there are disagreements about how to measure what matters to people (or indeed, whether it is conceptually possible to do so) and compare such measures to what people choose. [77] Values congruence should not be measured using the values clarity subscale of the Decisional Conflict Scale, as this subscale measures perceived values clarity, not values congruence. [48] Further research is required to determine whether measuring values congruence might introduce bias or otherwise negatively influence decision making.

Decisional regret should be assessed with a version of the Decisional Regret Scale [78, 79] after people make the decision, ideally with a sufficiently long delay that longer-term effects can be captured. An included study in this review showed that a values clarification method reduced decisional regret, but only after a year had passed following implementation of the decision. [54]

For all four measures, authors should clearly report sample mean and sample standard deviation for continuous measures, numbers in each category for categorical measures, and sample size per study arm in all cases. Finally, we recommend that patient decision aid developers explain the rationale for their choice of values clarification method.

Our study has three main limitations. First, the included data were of moderate quality. Although this review includes many robust trials, the included studies often measured different outcomes or the same outcomes in different ways, there were missing data in some studies, some studies had high risk of bias (often because it was not possible to prevent study participants from ascertaining the study arm to which they were assigned), and some of our meta-analyses had high heterogeneity. Together, these issues suggest a degree of caution in our conclusions. Second, we did not distinguish between subtypes of values clarification methods. For example, different adaptive conjoint analysis exercises may be very different from each other, as might open discussions, or many other values clarification methods we grouped together, particularly those we grouped under the broad umbrella term of multicriteria decision analysis. Indeed, the values clarification methods used and trialed may simply reflect authors’ interests and expertise. Given the breadth of methods available, further comparative effectiveness research is needed to conclusively determine the superiority of any given method. Third and finally, our primary findings were heavily influenced by studies conducted with relatively homogenous populations making hypothetical decisions. Although our sensitivity analyses suggested no differences between studies in real and hypothetical contexts, we nonetheless believe further study is needed in more diverse populations making real decisions before drawing firmer conclusions.

Our study also has three main strengths. First, we catalog definitions and resources regarding values clarification methods, as well as recommended outcomes to report in studies. In doing so, we hope to offer more clarity and structure to a literature that can be confusing to navigate, particularly for those who are newer to developing patient decision aids. Second, we begin to answer a core question that commonly arises when developing a patient decision aid: when including a values clarification method, which type of method should one use? Third and finally, we used rigorous methods and an expansive, systematic search. By conducting a systematic review, we reduced our likelihood of missing relevant studies. By including meta-analyses, we offer stronger findings and recommendations than would be possible without pooling data across multiple studies.

In conclusion, patient decision aids should include an explicit values clarification method. Patient decision aid developers may wish to consider the potential advantages of multicriteria decision analysis. Future research should further investigate which methods lead to the best outcomes across or within particular decisions, populations, and settings. Authors of randomized controlled trials in this area should report decisional conflict, decision made, values congruence, and decisional regret.

## Data Availability

Full data are available in the appendices.

## DECLARATIONS

### Abbreviations

None

### Ethics Approval, Consent to Participate and Consent for Publication

Not applicable.

### Availability of Data and Materials

Full data are available in the online appendices.

### Competing Interests

None.

### Funding

This study was funded by the Canadian Institutes of Health Research (CIHR) FDN-148426 (PI Witteman). HOW receives salary support from Tier 2 Canada Research Chair in Human-Centred Digital Health and received salary support during this study from a Fonds de Recherche du Québec-Santé (FRQS) Research Scholar Junior 2 Career Award. The CIHR, Canada Research Chairs program, and FRQS had no role in determining the study design, the plans for data collection or analysis, the decision to publish, nor the preparation of this manuscript.

### Authors’ Contributions

All authors contributed to the design of the study. HOW, RN, GV, SCD, SC, MM, TP, CR, CRB, JJ contributed to data collection. HOW, SCD, and JJ conducted data analysis and interpretation. HOW drafted the first version of the article with early revision by RN, GV, SCD, SC, MM, TP, CR, CRB, JJ and multiple subsequent revisions by all authors. All authors critically revised the article and approved the final version for submission for publication. HOW had full access to all the data in the study and had final responsibility for the decision to submit for publication. Authors BA, JFPB, SC, AF, TG, MM, AP, MP, TP, CR, DR, CRB, PT, MW, DBW, CEW contributed approximately equally and are listed alphabetically by last name.

## Acknowledgements

The authors gratefully acknowledge Frédéric Bergeron, MLIS, for assistance with search strategy and Caroline Beaudoin for assistance in resolving article counts. We thank all authors of the original articles who generously gave their time to provide missing data when we were unable to extract the data needed from their papers.

## Online Appendix 1: Search Strategies

**#1** ((value? OR patient preference? OR treatment preference?) adj5 (clarif* OR elicit*)).ti,ab,kw

**#2** (Decision Making/ AND Social Values/)

**#3** MCDA.ti,ab

**#4** analytical hierarchy process.ti,ab,kw

**#5** best-worst scaling.ti,ab,kw

**#6** ((conjoint OR decision) adj3 analysis):ti,ab,kw

**#7** data envelopment analysis.ti,ab,kw

**#8** Decision conferencing.ti,ab,kw

**#9** Decision models.ti,ab,kw

**#10** direct rating.ti,ab,kw

**#11** points allocation.ti,ab,kw

**#12** discrete choice experiment.ti,ab, kw

**#13** (“dominance-based” adj3 approach*).ti,ab,kw

**#14** EVIDEM framework.ti,ab,kw

**#15** (geometrical analysis for interactive aid OR GAIA).ti,ab,kw

**#16** MACBETH.ti,ab,kw

**#17** (“Measuring Attractiveness” adj4 “Categorical Based Evaluation TecHnique”).ti,ab,kw

**#18** Multi-Attribute Global Inference of Quality.ti,ab,kw

**#19** ((“Multiple attribute” OR “multiple criteria” OR multiattribute) adj2 (utility OR theory OR analysis)).ti,ab,kw

**#20** (MAUT OR MAVT OR MCUA OR MCA).ti,ab,kw

**#21** (“Novel approach to imprecise assessment and decision environments” OR NAIADE).ti,ab,kw

**#22** ORESTE.ti,ab,kw

**#23** Pairwise comparisons.ti,ab,kw

**#24** PAPRIKA.ti,ab,kw

**#25** Pairwise RanKings.ti,ab,kw

**#26** PROMETHEE.ti,ab,kw

**#27** Preference Ranking Organization Method for Enrichment of Evaluations.ti,ab,kw

**#28** QUALItative FLEXible.ti,ab,kw

**#29** Simple Multi Attribute Rating Technique.ti,ab,kw

**#30** SMART.ti,ab,kw

**#31** Standard gamble.ti,ab,kw

**#32** Swing weighting.ti,ab,kw

**#33** TOPSIS.ti,ab,kw

**#34** Technique for Order Preference by Similarity to the Ideal Solution.ti,ab,kw

**#35** (Time tradeoff OR time tradeoff).ti,ab, kw

**#36** Value function methods.ti,ab,kw

**#37** Valutazione delle Tecnologie Sanitarie.ti,ab,kw

**#38** VDA.ti,ab,kw

**#39** VTS.ti,ab,kw

**#40** verbal decision analysis.ti,ab,kw

**#41** visual analog scale.ti,ab,kw

**#42** willingness-to-pay.ti,ab,kw

**#43** ((Scoring OR weighting) adj1 methods).ti,ab,kw

**#44** REGIME.ti,ab,kw

**#45** (scal* adj2 (methods OR Natural OR Constructed OR Objective)).ti,ab,kw

**#46** OR/3-45

**#47** Randomized Controlled Trials as Topic/

**#48** randomized controlled trial/

**#49** Random Allocation/

**#50** Double Blind Method/

**#51** Single Blind Method/

**#52** clinical trial/

**#53** clinical trial, phase i.pt

**#54** clinical trial, phase ii.pt

**#55** clinical trial, phase iii.pt

**#56** clinical trial, phase iv.pt

**#57** controlled clinical trial.pt

**#58** randomized controlled trial.pt

**#59** multicenter study.pt

**#60** clinical trial.pt

**#61** exp Clinical Trials as topic/

**#62** (clinical adj trial$).tw

**#63** ((singl$ or doubl$ or treb$ or tripl$) adj (blind$3 or mask$3)).tw

**#64** PLACEBOS/

**#65** placebo$.tw

**#66** randomly allocated.tw

**#67** (allocated adj2 random$).tw

**#68** OR/47-67

**#69** (#1 OR #2 OR #46) AND #68

**#70** #70 NOT (animals/ NOT humans/)

**#1** ((value* OR “patient preference” OR “treatment preferences”) NEAR/5 (clarif* OR elicit*)):ti,ab,kw

**#2** ‘decision support system’/de OR ‘patient decision making’/de

**#3** MCDA:ti,ab,kw

**#4** “analytical hierarchy process”:ti,ab,kw

**#5** “best-worst scaling”:ti,ab,kw

**#6** ((conjoint OR decision) NEAR/3 analysis):ti,ab,kw

**#7** “data envelopment analysis”:ti,ab,kw

**#8** “Decision conferencing”:ti,ab,kw

**#9** “Decision models”:ti,ab,kw

**#10** “direct rating”:ti,ab,kw

**#11** “points allocation”:ti,ab,kw

**#12** “discrete choice experiment”:ti,ab,kw

**#13** (“dominance-based” NEAR/3 approach*):ti,ab,kw

**#14** “EVIDEM framework”:ti,ab,kw

**#15** “geometrical analysis for interactive aid OR GAIA”:ti,ab,kw

**#16** MACBETH:ti,ab,kw

**#17** (“Measuring Attractiveness” NEAR/4 “Categorical Based Evaluation TecHnique”):ti,ab,kw

**#18** “Multi-Attribute Global Inference of Quality”:ti,ab,kw

**#19** ((“Multiple attribute” OR “multiple criteria” OR multiattribute) NEAR/2 (utility OR theory OR analysis)):ti,ab,kw

**#20** (MAUT OR MAVT OR MCUA OR MCA):ti,ab,kw

**#21** (“Novel approach to imprecise assessment and decision environments” OR NAIADE):ti,ab,kw

**#22** ORESTE:ti,ab,kw

**#23** “Pairwise comparisons”:ti,ab,kw

**#24** PAPRIKA:ti,ab,kw

**#25** “Pairwise RanKings”:ti,ab,kw

**#26** PROMETHEE:ti,ab,kw

**#27** “Preference Ranking Organization Method for Enrichment of Evaluations”:ti,ab,kw

**#28** “QUALItative FLEXible”:ti,ab,kw

**#29** “Simple Multi Attribute Rating Technique”:ti,ab,kw

**#30** SMART:ti,ab,kw

**#31** “Standard gamble”:ti,ab,kw

**#32** “Swing weighting”:ti,ab,kw

**#33** TOPSIS:ti,ab,kw

**#34** “Technique for Order Preference by Similarity to the Ideal Solution”:ti,ab,kw

**#35** (“Time tradeoff” OR “time tradeoff”):ti,ab,kw

**#36** “Value function methods”:ti,ab,kw

**#37** “Valutazione delle Tecnologie Sanitarie”:ti,ab,kw

**#38** VDA:ti,ab,kw

**#39** VTS:ti,ab,kw

**#40** “verbal decision analysis”:ti,ab,kw

**#41** “visual analog scale”:ti,ab,kw

**#42** “willingness-to-pay”:ti,ab,kw

**#43** ((Scoring OR weighting) NEAR/1 methods):ti,ab,kw

**#44** REGIME:ti,ab,kw

**#45** (scal* NEAR/2 (methods OR Natural OR Constructed OR Objective)):ti,ab,kw

**#46** #3 OR #4 OR #5 OR #6 OR #7 OR #8 OR #9 OR #10 OR #11 OR #12 OR #13 OR #14 OR #15 OR #16 OR #17 OR #18 OR #19 OR #20 OR

**#47** ‘clinical trial’/de

**#48** ‘randomized controlled trial’/de

**#49** ‘randomization’/de

**#50** ‘single blind procedure’/de

**#51** ‘double blind procedure’/de

**#52** ‘crossover procedure’/de

**#53** ‘placebo’/de OR ‘prospective study’/de

**#54** (randomi?ed NEXT/1 controlled NEXT/1 trial):ab,ti

**#55** rct:ab,ti OR ‘random allocation’:ab,ti

**#56** ‘randomly allocated’:ab,ti OR ‘allocated randomly’:ab,ti

**#57** (allocated NEAR/2 random):ab,ti

**#58** ((single OR double OR treble OR triple) NEXT/1 blind*):ab,ti

**#59** placebo*:ab,ti

**#60** #47 OR #48 OR #49 OR #50 OR #51 OR #52 OR #53 OR #54 OR #55 OR #56 OR #57 OR #58 OR #59

**#61** (#1 OR #2 OR #46) AND #60

**#62** #61 NOT (animal:de NOT human:de)

**#63** [embase]/lim NOT ([embase]/lim AND [medline]/lim)

**#1** ((value* OR “patient preference*” OR “treatment preference*”) NEAR/5 (clarif* OR elicit*)):ti,ab

**#2** (“Decision Making”:kw AND “Social Values”:kw)

**#3** MCDA:ti,ab

**#4** “analytical hierarchy process”:ti,ab

**#5** best-worst scaling:ti,ab

**#6** ((conjoint OR decision) NEAR/3 analysis):ti,ab

**#7** “Decision conferencing”:ti,ab

**#8** “Decision models”:ti,ab

**#9** “direct rating”:ti,ab

**#10** “points allocation”:ti,ab

**#11** “discrete choice experiment”:ti,ab

**#12** (“dominance-based” NEAR/3 approach*):ti,ab

**#13** “Elimination and Choice Expressing Reality”:ti,ab

**#14** “EVIDEM framework”:ti,ab

**#15** “geometrical analysis for interactive aid” OR GAIA:ti,ab

**#16** MACBETH:ti,ab

**#17** (“Measuring Attractiveness” NEAR/4 “Categorical Based Evaluation TecHnique”):ti,ab

**#18** “Multi-Attribute Global Inference of Quality”:ti,ab

**#19** “multiattribute objective function specification”:ti,ab

**#20** ((“Multiple attribute” OR “multiple criteria” OR multiattribute) NEAR/2 (utility OR theory OR analysis)):ti,ab

**#21** (MAUT OR MAVT OR MCUA OR MCA):ti,ab

**#22** (“Novel approach to imprecise assessment and decision environments” OR NAIADE):ti,ab

**#23** ORESTE:ti,ab

**#24** “Pairwise comparisons”:ti,ab

**#25** PAPRIKA:ti,ab

**#26** “Pairwise RanKings”:ti,ab

**#27** PROMETHEE:ti,ab

**#28** “Preference Ranking Organization Method for Enrichment of Evaluations”:ti,ab

**#29** “QUALItative FLEXible”:ti,ab

**#30** “Simple Multi Attribute Rating Technique”:ti,ab

**#31** SMART:ti,ab

**#32** “Standard gamble”:ti,ab

**#33** “Swing weighting”:ti,ab

**#34** TOPSIS:ti,ab

**#35** “Technique for Order Preference by Similarity to the Ideal Solution”:ti,ab

**#36** (“Time tradeoff” OR “time tradeoff”):ti,ab

**#37** “Value function methods”:ti,ab

**#38** “Valutazione delle Tecnologie Sanitarie”:ti,ab

**#39** VDA:ti,ab

**#40** VTS:ti,ab

**#41** “verbal decision analysis”:ti,ab

**#42** “visual analog scale”:ti,ab

**#43** “willingness-to-pay”:ti,ab

**#44** scoring method*:ti,ab OR “weighting method*”:ti,ab

**#45** REGIME:ti,ab

**#46** (scal* NEAR/2 (methods OR Natural OR Constructed OR Objective)):ti,ab

**#47** {OR #3-#46}

**#48** #1 OR #2 OR #47

**#1** TS=(((value$ OR “patient preference$” OR “treatment preference$”) NEAR/5 (clarif* OR elicit*)))

**#2** TS=((conjoint OR decision) NEAR/3 analysis)

**#3** TS=(MCDA)

**#4** TS=(“analytical hierarchy process”)

**#5** TS=(“best-worst scaling”)

**#6** TS=(“data envelopment analysis”)

**#7** TS=(“Decision conferencing”)

**#8** TS=(“Decision models”)

**#9** TS=(“direct rating”)

**#10** TS=(“points allocation”)

**#11** TS=(“discrete choice experiment”)

**#12** TS=(“dominance-based” NEAR/3 approach*)

**#13** TS=(“Elimination and Choice Expressing Reality”) **#14** TS=(“EVIDEM framework”)

**#15** TS=(“geometrical analysis for interactive aid” OR GAIA)

**#16** TS=(MACBETH)

**#17** TS= (“Measuring Attractiveness” NEAR/4 “Categorical Based Evaluation TecHnique”)

**#18** TS=(“Multi-Attribute Global Inference of Quality”)

**#19** TS=((“Multiple attribute” OR “multiple criteria” OR multiattribute) NEAR/2 (utility OR theory OR analysis))

**#20** TS=(MAUT OR MAVT OR MCUA OR MCA)

**#21** TS=(“Novel approach to imprecise assessment and decision environments” OR NAIADE)

**#22** TS=(ORESTE)

**#23** TS=(“Pairwise comparisons”)

**#24** TS=(PAPRIKA)

**#25** TS=(“Pairwise RanKings”)

**#26** TS=(PROMETHEE)

**#27** TS=(“Preference Ranking Organization Method for Enrichment of Evaluations”)

**#28** TS=(“QUALItative FLEXible”)

**#29** TS=(“Simple MultiAttribute Rating Technique”)

**#30** TS=(SMART)

**#31** TS=(“Standard gamble”)

**#32** TS=(TOPSIS)

**#33** TS=(“Technique for Order Preference by Similarity to the Ideal Solution”)

**#34** TS=(“time tradeoff” OR “time tradeoff”)

**#35** TS=(“Value function methods”)

**#36** TS=(“Valutazione delle Tecnologie Sanitarie”)

**#37** TS=(VDA)

**#38** TS=(VTS)

**#39** TS=(“verbal decision analysis”)

**#40** TS=(“visual analog scale”)

**#41** TS=(“willingness-to-pay”)

**#42** TS= ((Scoring OR weighinng) NEAR/1 methods)

**#43** TS=(REGIME)

**#44** TS=(scal* NEAR/2 (methods OR Natural OR Constructed OR Objective))

#44 OR #43 OR #42 OR #41 OR #40 OR #39 OR #38 OR #37 OR #36 OR #35 OR #34 OR #33 OR #32 OR #31 OR #30 OR #29 OR #28 OR

#27 OR #26 OR #25 OR #24 OR #23 OR #22 OR #21 OR #20 OR #19 OR #18 OR #17 OR #16 OR #15 OR #14 OR #13 OR #12 OR #11 OR

**#45** #10 OR #9 OR #8 OR #7 OR #6 OR #5 OR #4 OR #3 OR #2

**#46** “clinical trial”

**#47** randomization

**#48** “crossover procedure”

**#49** placebo

**#50** “prospective study”

**#51** (randomi$ed NEAR/1 “controlled trial”)

**#52** rct

**#53** (allocat* NEAR/2 random*)

**#54** ((single OR double OR treble OR triple) NEAR/1 blind*))

**#55** #46 OR #47 OR #48 OR #49 OR #50 OR #51 OR #52 OR #53 #54

**#56** (#1 OR #45) AND #55

**#1** TI ((value# OR “patient preference#” OR “treatment preference#”) N5 ((clarif* OR elicit*))

**#2** AB ((value# OR “patient preference#” OR “treatment preference#”) N5 ((clarif* OR elicit*))

**#3** MH “Values Clarification”

**#4** MM “Decision Support Techniques”

**#5** TI ((conjoint OR decision) N3 analysis) OR AB ((conjoint OR decision) N3 analysis)

**#6** TI MCDA OR AB MCDA

**#7** TI “analytical hierarchy process” OR AB “analytical hierarchy process”

**#8** TI “best-worst scaling” OR AB “best-worst scaling”

**#9** TI “data envelopment analysis” OR AB “data envelopment analysis”

**#10** TI “Decision conferencing” OR AB “Decision conferencing”

**#11** TI “Decision models” OR AB “Decision models”

**#12** TI “direct rating” OR AB “direct rating”

**#13** TI “points allocation” OR AB “points allocation”

**#14** TI “discrete choice experiment” OR AB “discrete choice experiment”

**#15** TI (“dominance-based” N3 approach*) OR AB (“dominance-based” N3 approach*)

**#16** TI “Elimination and Choice Expressing Reality” OR AB “Elimination and Choice Expressing Reality”

**#17** TI “EVIDEM framework” OR AB “EVIDEM framework”

**#18** TI “geometrical analysis for interactive aid” OR GAIA OR AB “geometrical analysis for interactive aid” OR GAIA

**#19** TI MACBETH OR AB MACBETH

**#20** TI (“Measuring Attractiveness” N4 “Categorical Based Evaluation TecHnique”) OR AB (“Measuring Attractiveness” N4 “

**#21** TI ((“Multiple attribute” OR “multiple criteria” OR multiattribute) N2 (utility OR theory OR analysis)) OR AB ((“Multiple

**#22** TI (MAUT OR MAVT OR MCUA OR MCA) OR AB (MAUT OR MAVT OR MCUA OR MCA)

**#23** TI ORESTE OR AB ORESTE

**#24** TI “Pairwise comparisons” OR AB “Pairwise comparisons”

**#25** TI PAPRIKA OR AB PAPRIKA

**#26** TI “Pairwise RanKings” OR AB “Pairwise RanKings”

**#27** TI SMART OR AB SMART

**#28** TI “Standard gamble” OR AB “Standard gamble”

**#29** TI “Swing weighting” OR AB “Swing weighting”

**#30** TI TOPSIS OR AB TOPSIS

**#31** TI “Technique for Order Preference by Similarity to the Ideal Solution” OR AB “Technique for Order Preference by Simi

**#32** TI (“Time tradeoff” OR “time tradeoff”) OR AB (“Time tradeoff” OR “time tradeoff”)

**#33** TI VDA OR AB VDA

**#34** TI VTS OR AB VTS

**#35** TI “visual analog scale” OR AB “visual analog scale”

**#36** TI “willingness-to-pay” OR AB “willingness-to-pay”

**#37** TI Scoring OR “weighting method#” OR AB Scoring OR “weighting method#”

**#38** TI REGIME OR AB REGIME

**#39** TI (scal* N2 (methods OR Natural OR Constructed OR Objective)) OR AB (scal* N2 (methods OR Natural OR Constructe

**#40** (S5 OR S6 OR S7 OR S8 OR S9 OR S10 OR S11 OR S12 OR S13 OR S14 OR S15 OR S16 OR S17 OR S18 OR S19 OR S20 OR

**#41** TX allocat* random*

**#42** MH “Quantitative Studies”

**#43** MH “Placebos”

**#44** TX placebo*

**#45** TX random* allocat*

**#46** MH “Random Assignment”

**#47** TX randomi* control* trial*

**#48** TX ((singl* n1 blind*) OR (singl* n1 mask*))

**#49** TX ((doubl* n1 blind*) OR (doubl* n1 mask*))

**#50** TX ((tripl* n1 blind*) OR (tripl* n1 mask*))

**#51** TX ((trebl* n1 blind*) or (trebl* n1 mask*))

**#52** TX (clinic* n1 trial*)

**#53** PT “Clinical trial”

**#54** MH “Clinical Trials+”

**#55** #52 OR #65

**#56** (#1 OR #2 OR #3 OR #4 OR #40) AND #55

## Online Appendix 2: Further Study Details

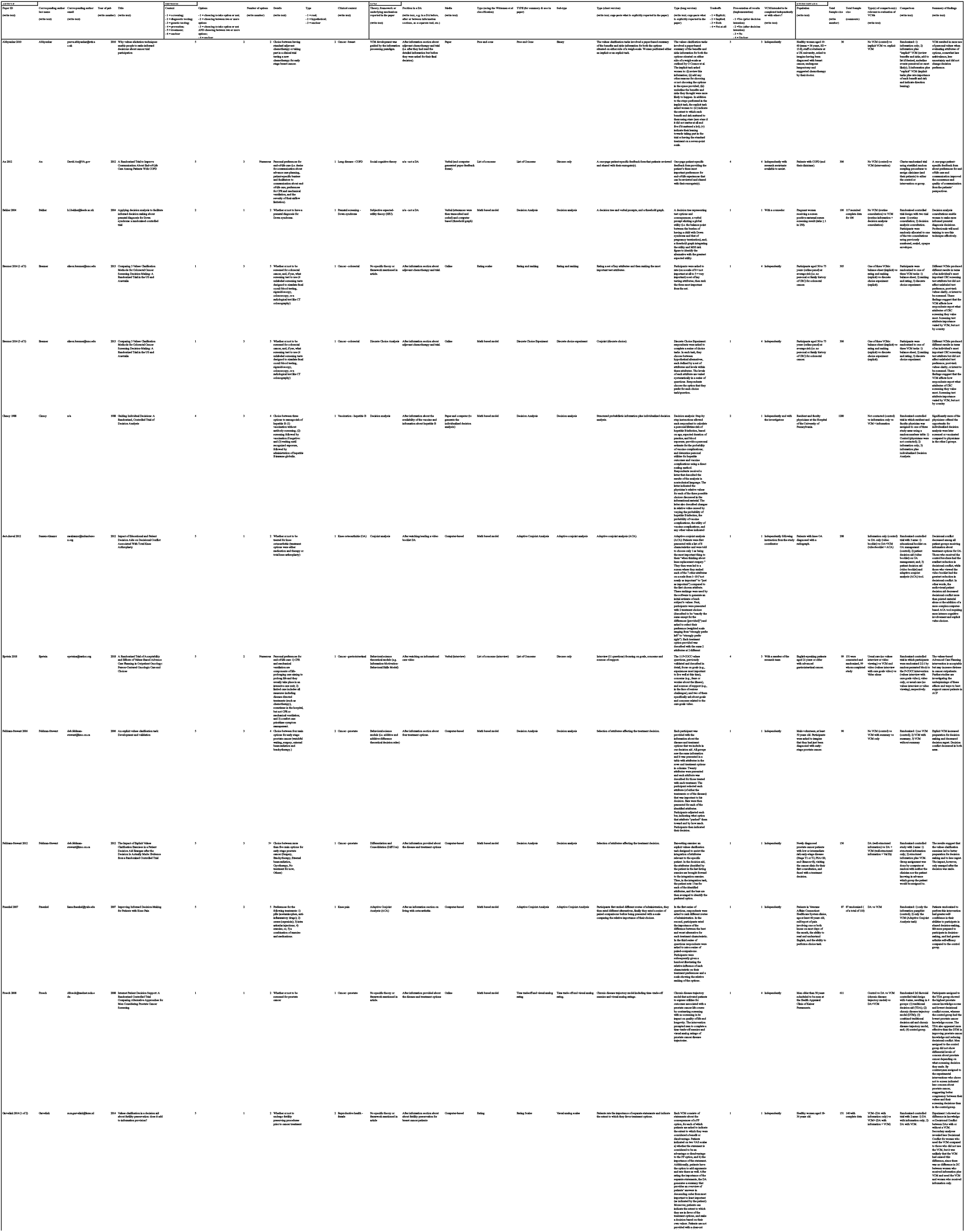
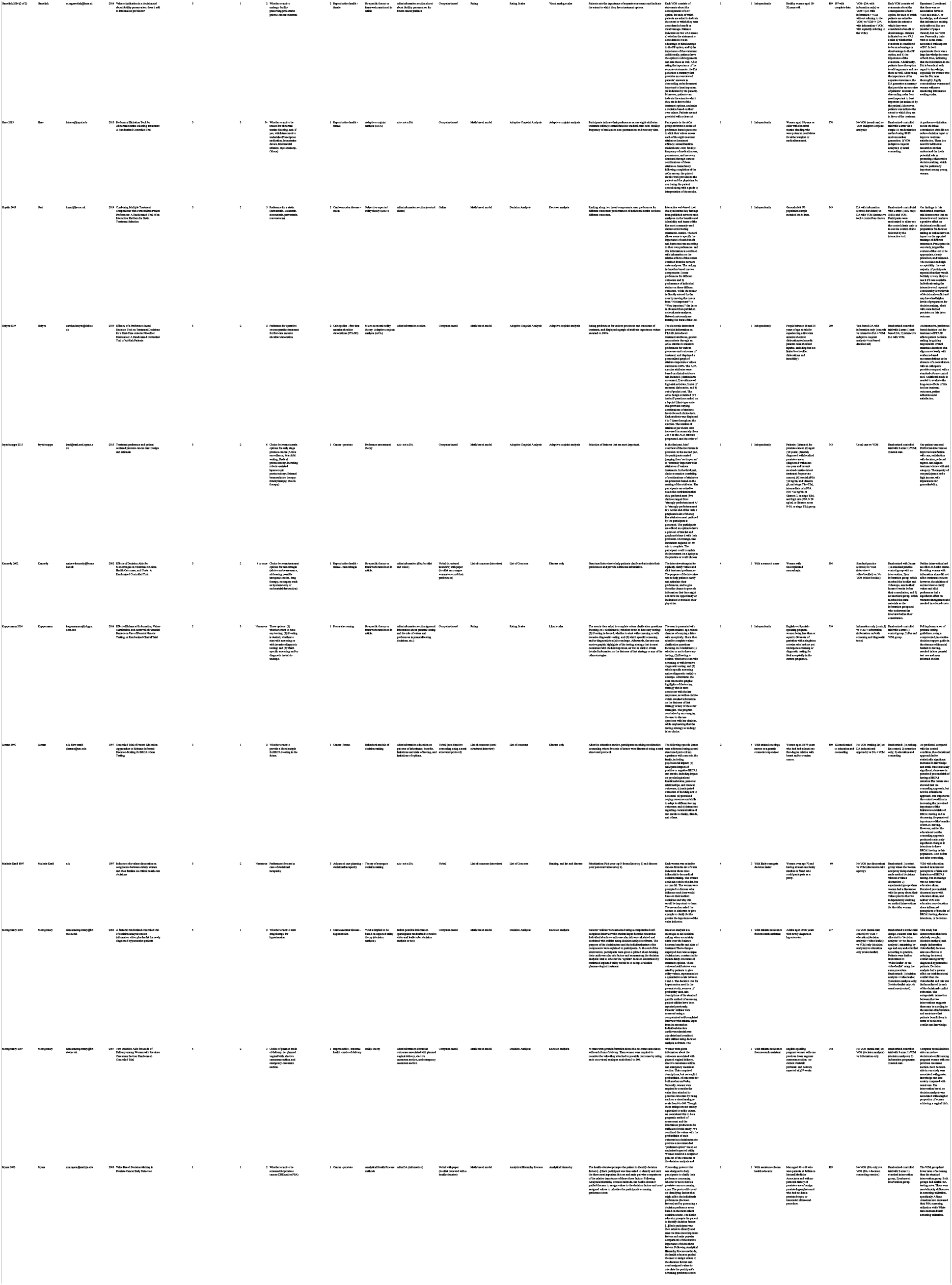
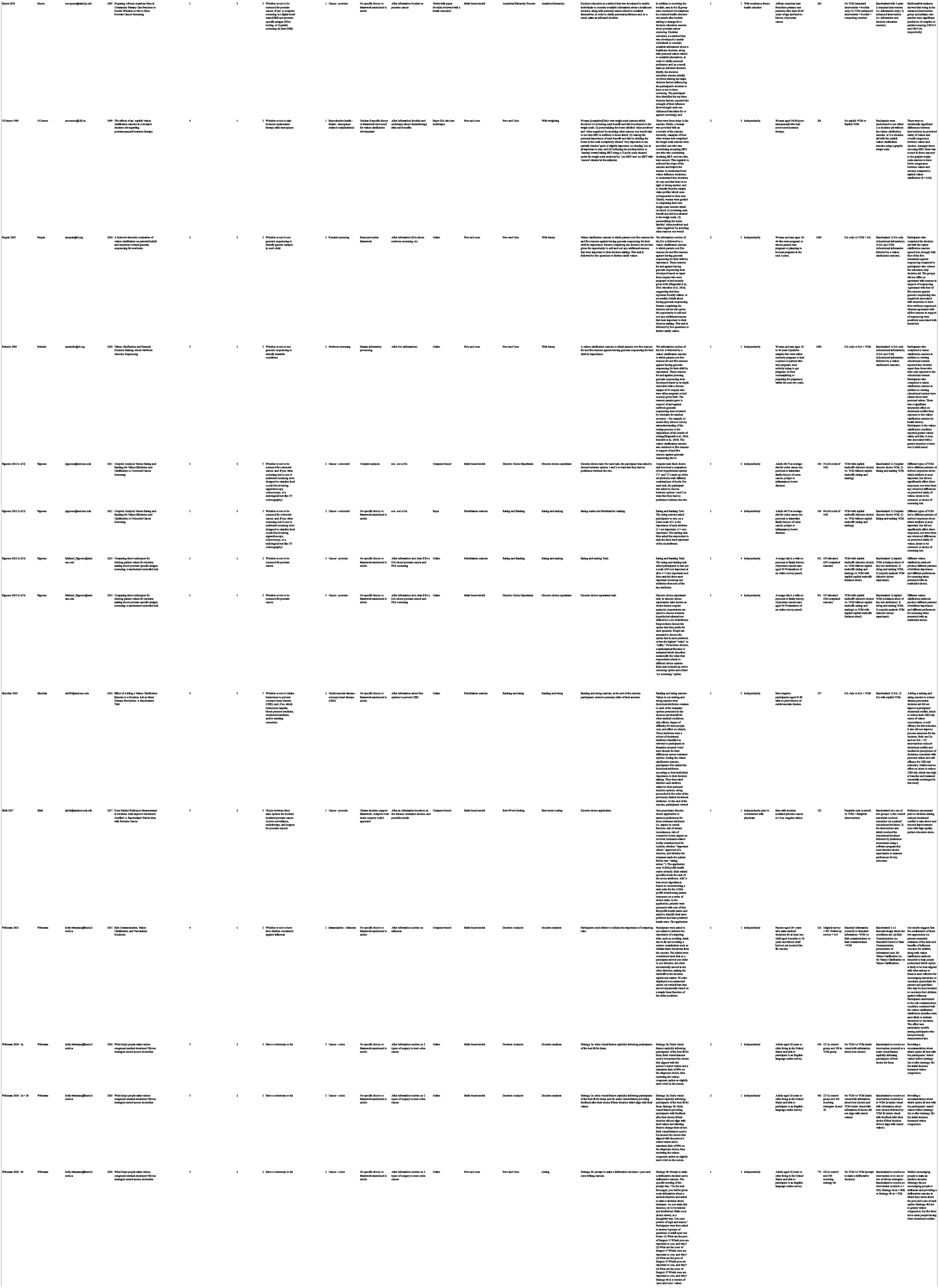
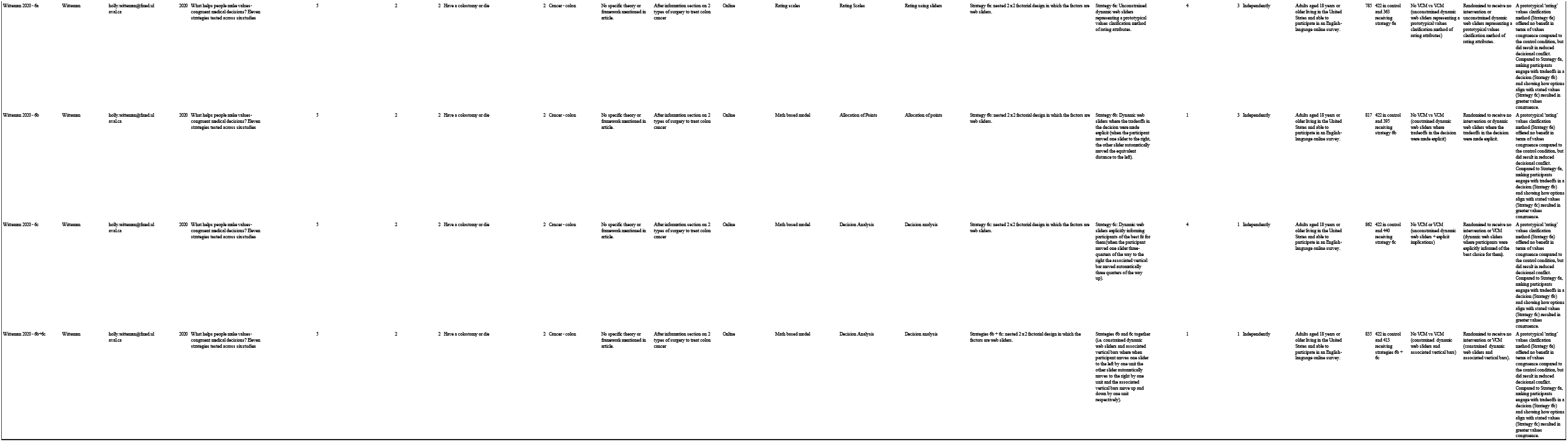

## Online Appendix 3. Additional Results

### Additional Meta-analytic Results

**Figure S1.**
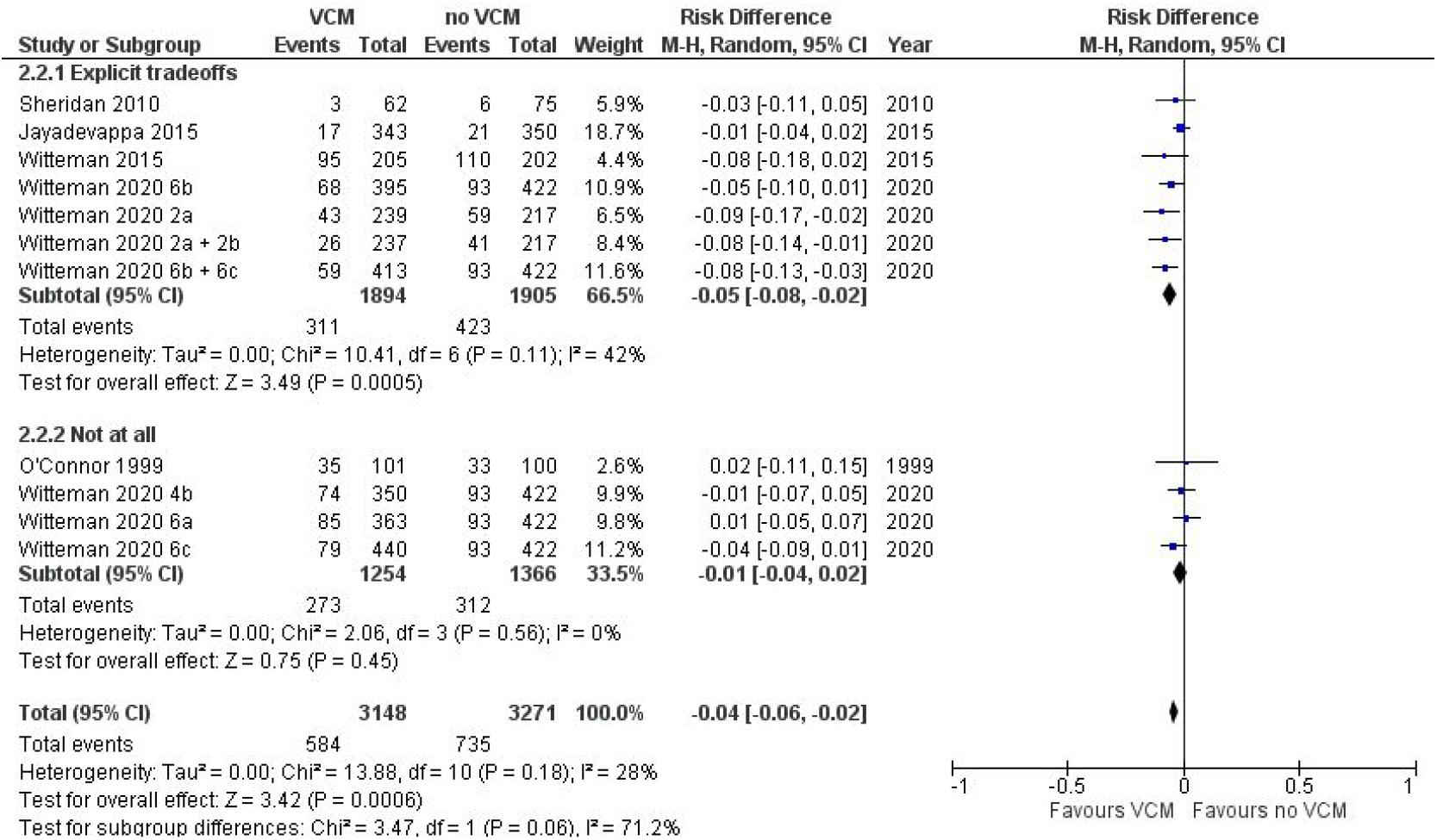
Values Congruence by Tradeoffs

**Figure S2.**
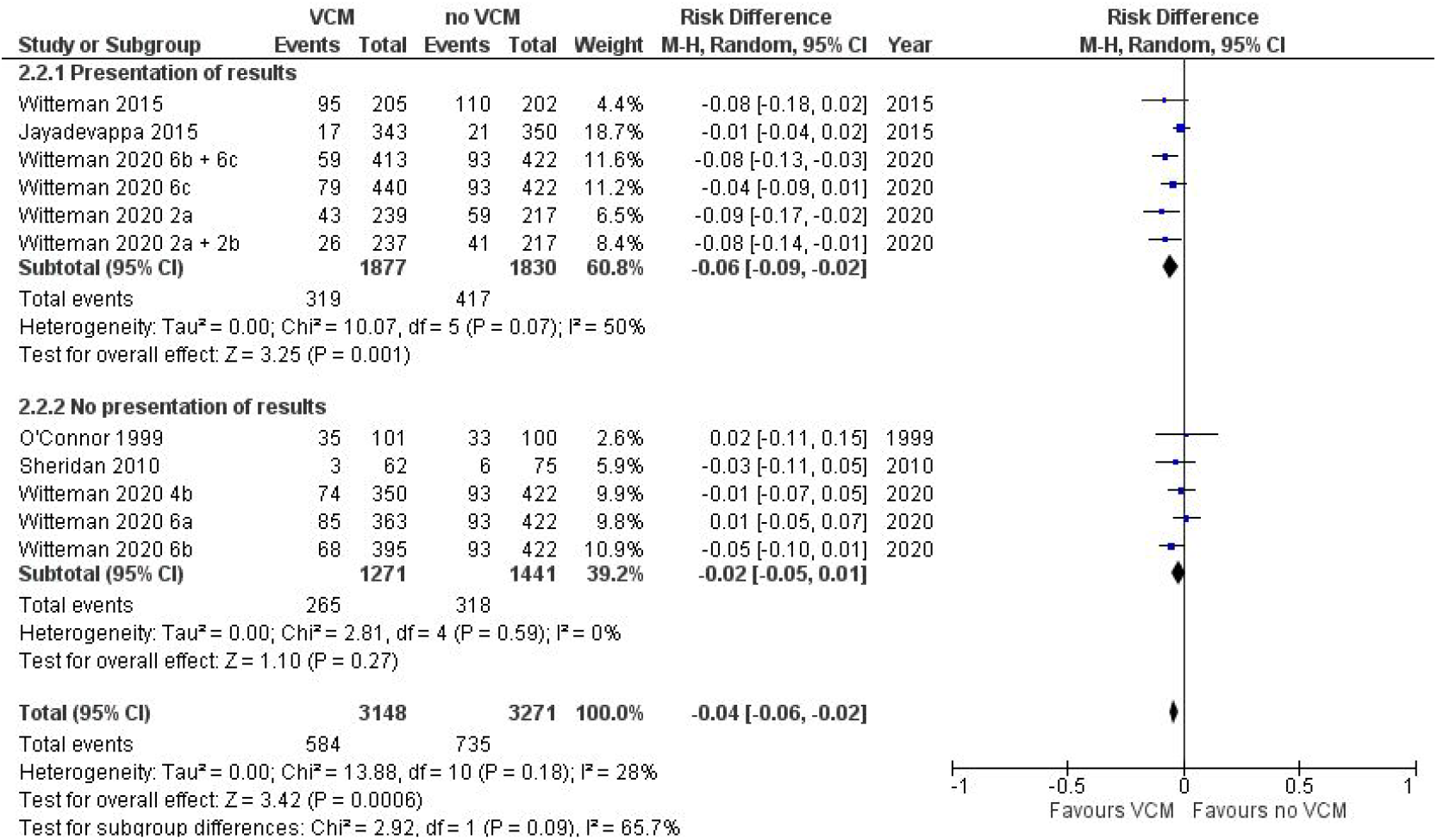
Values Congruence by Implementation/Presentation of Results

**Figure S3.**
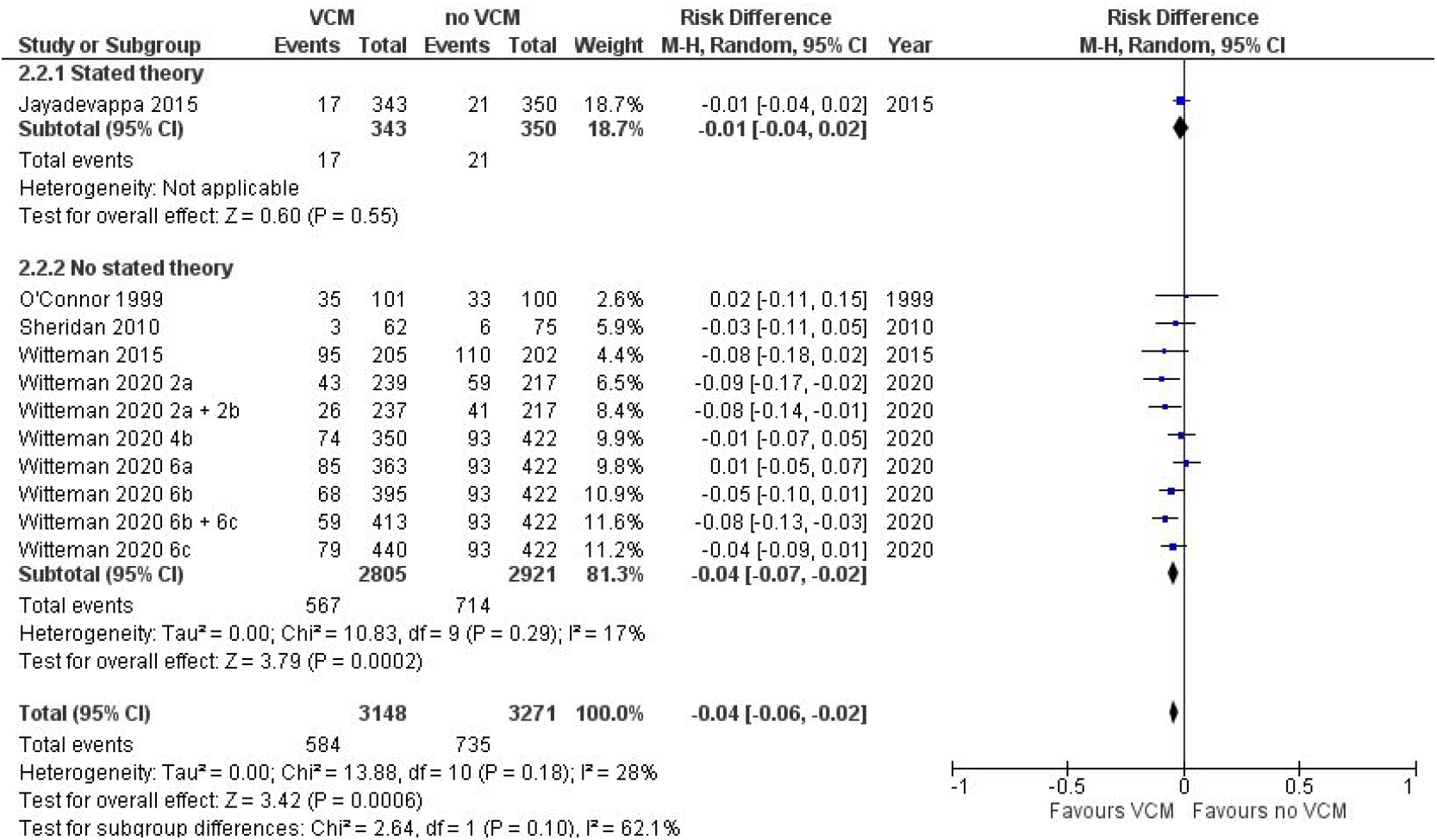
Values Congruence by Stated Use of a Theory/Framework

**Figure S4.**
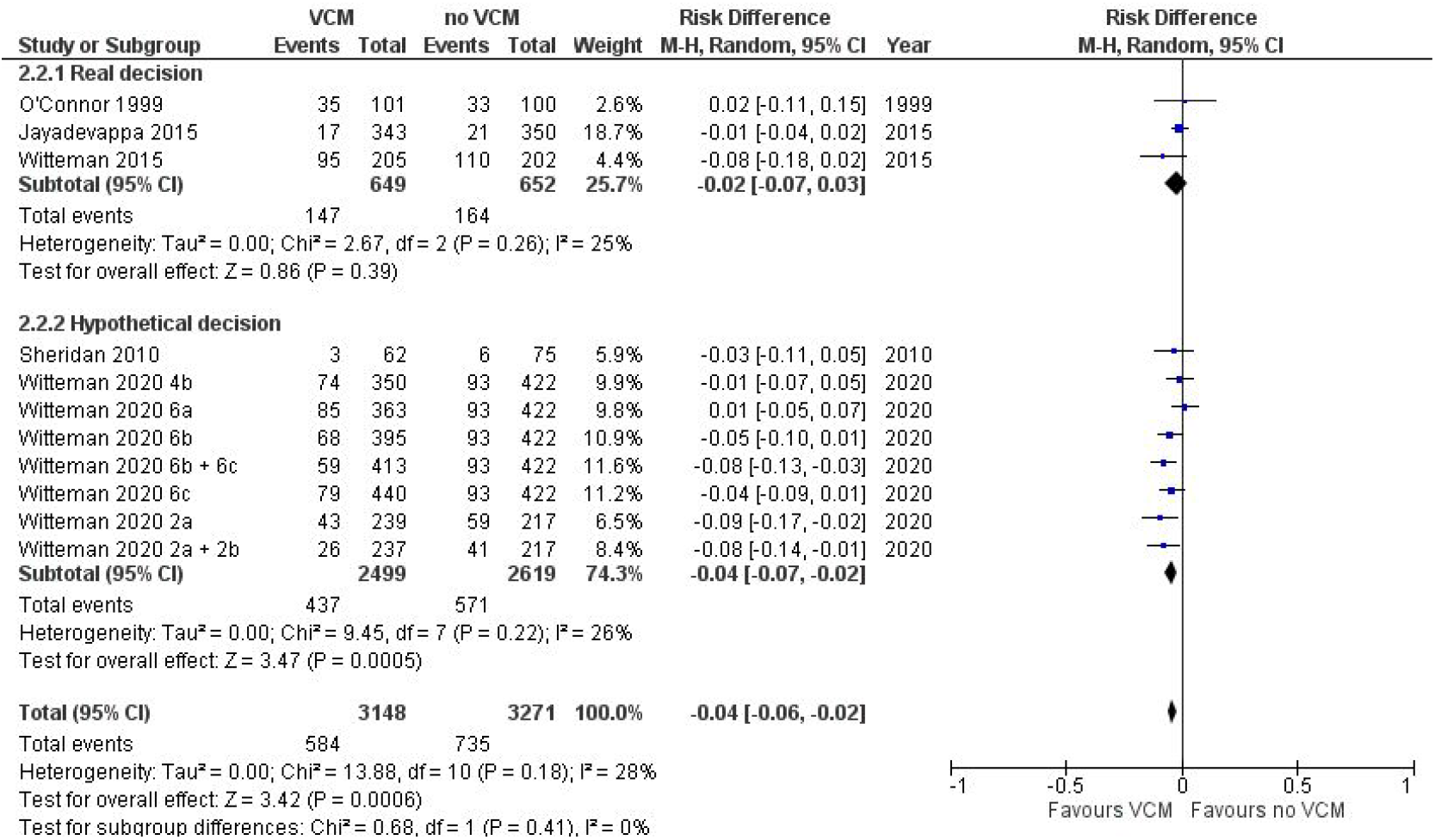
Values Congruence by Real/Hypothetical Decision

**Figure S5.**
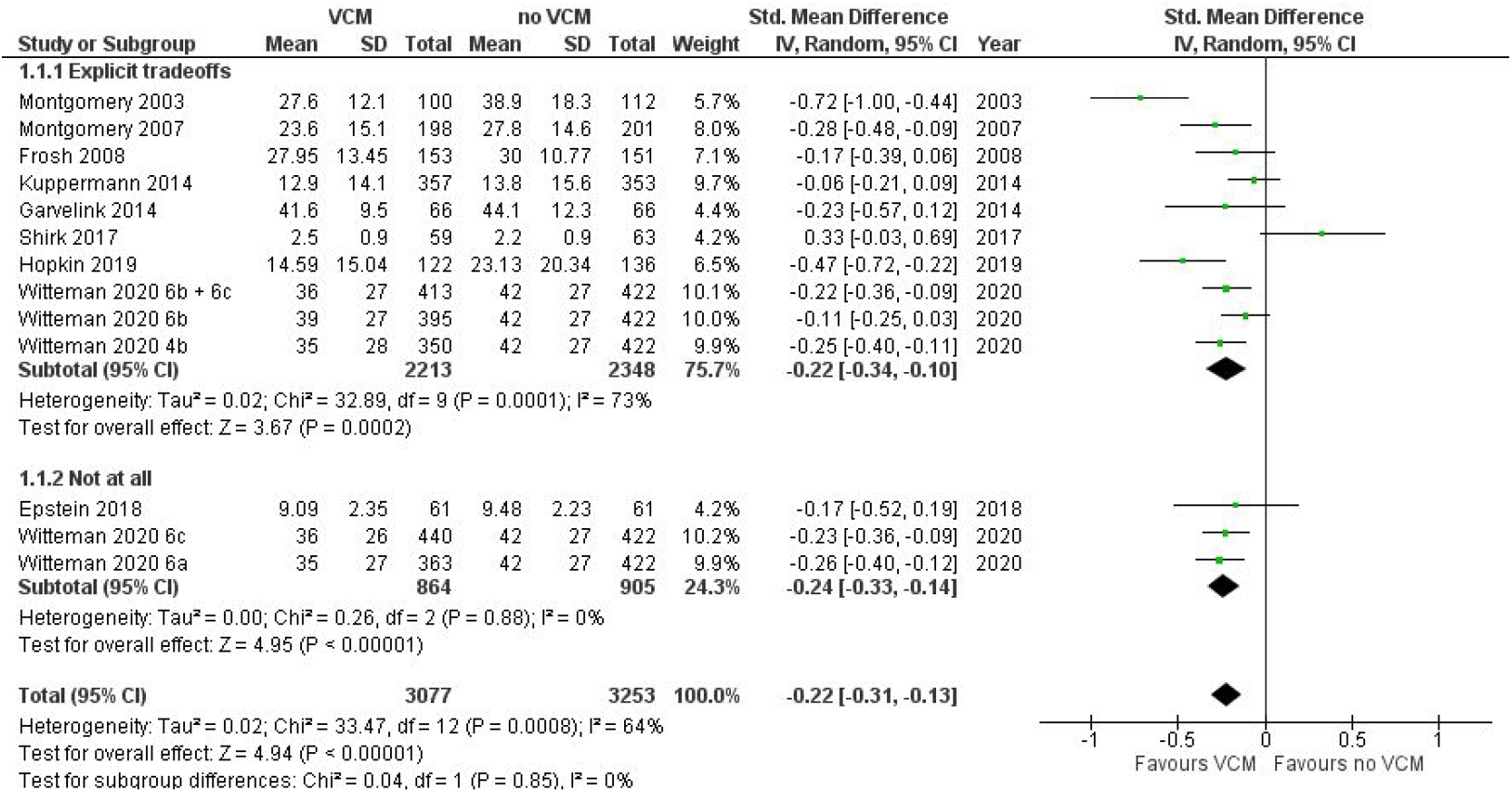
Decisional Conflict by Tradeoffs

**Figure S6.**
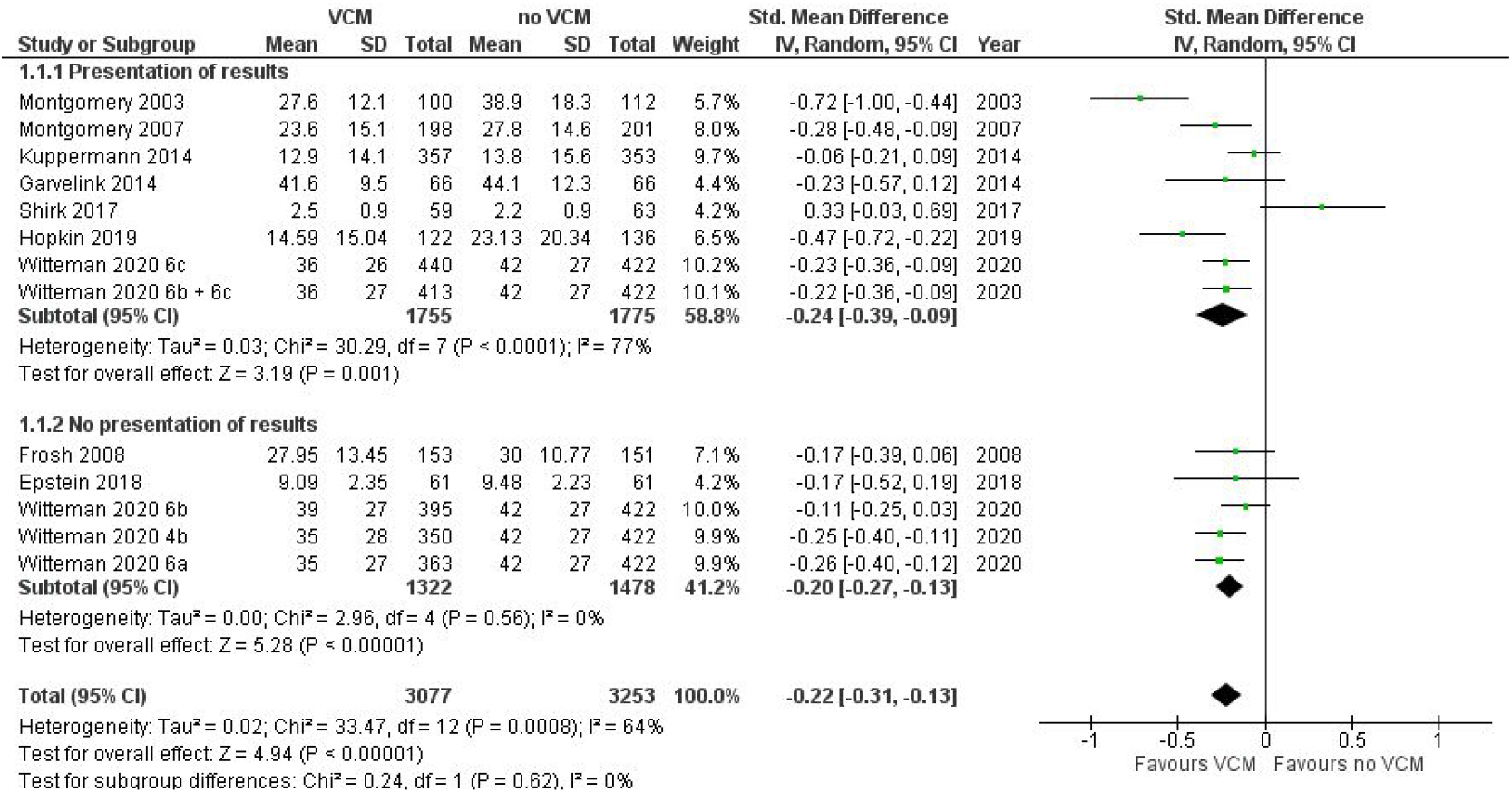
Decisional Conflict by Implications/Presentation of Results

**Figure S7.**
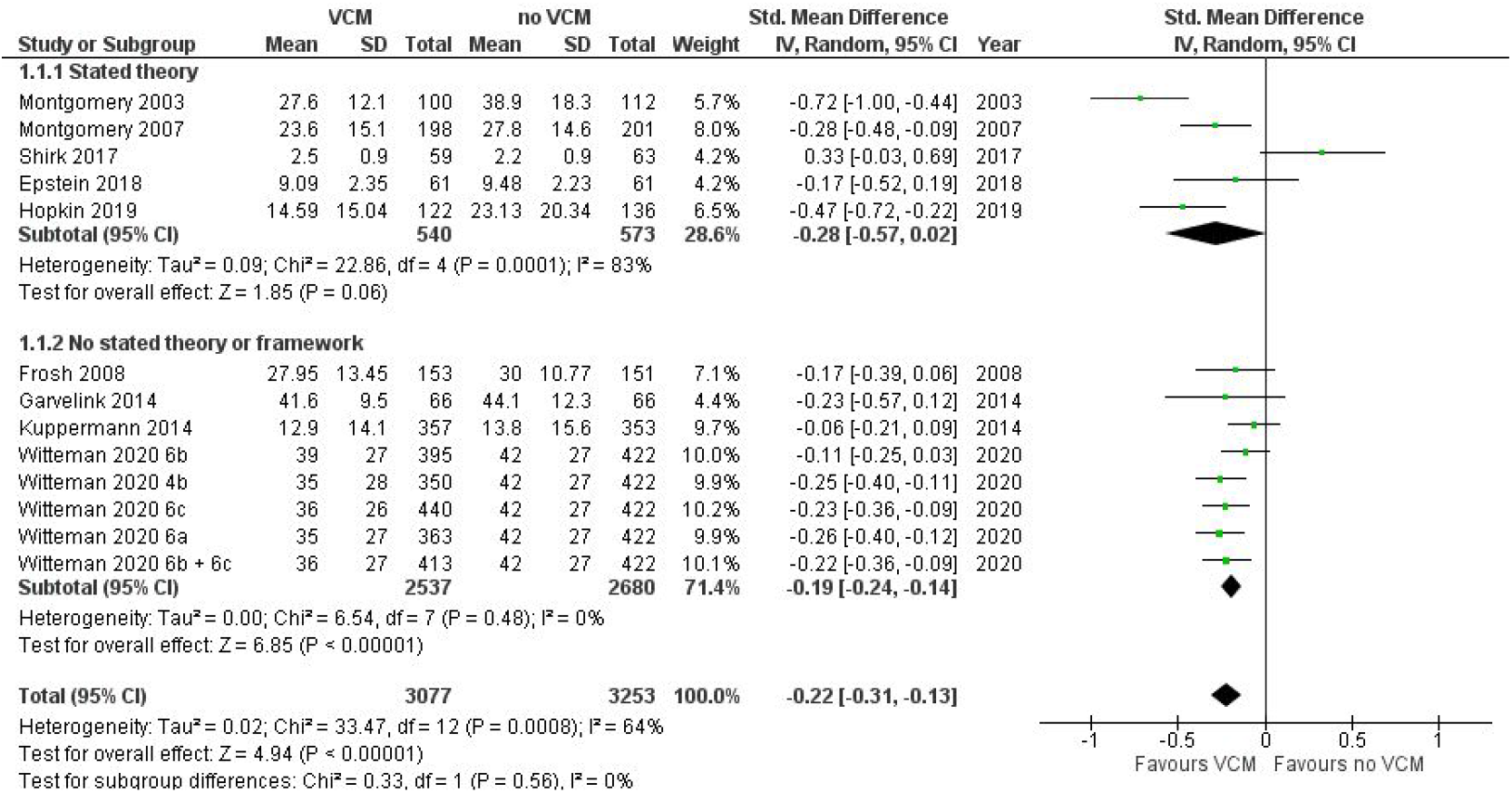
Decisional Conflict by Stated Use of a Theory/Framework

**Figure S8.**
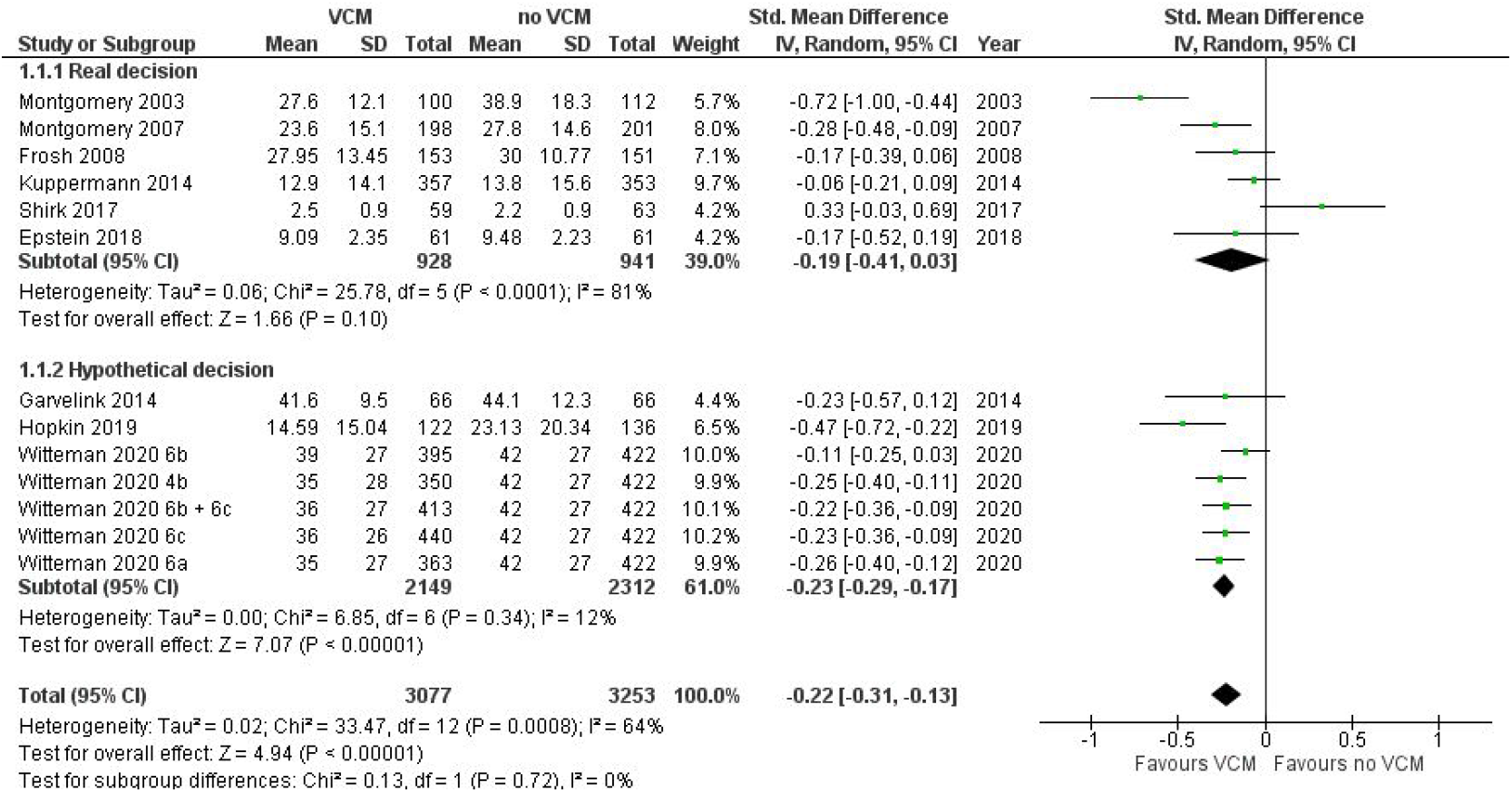
Decisional Conflict by Real/Hypothetical

**Figure S9.**
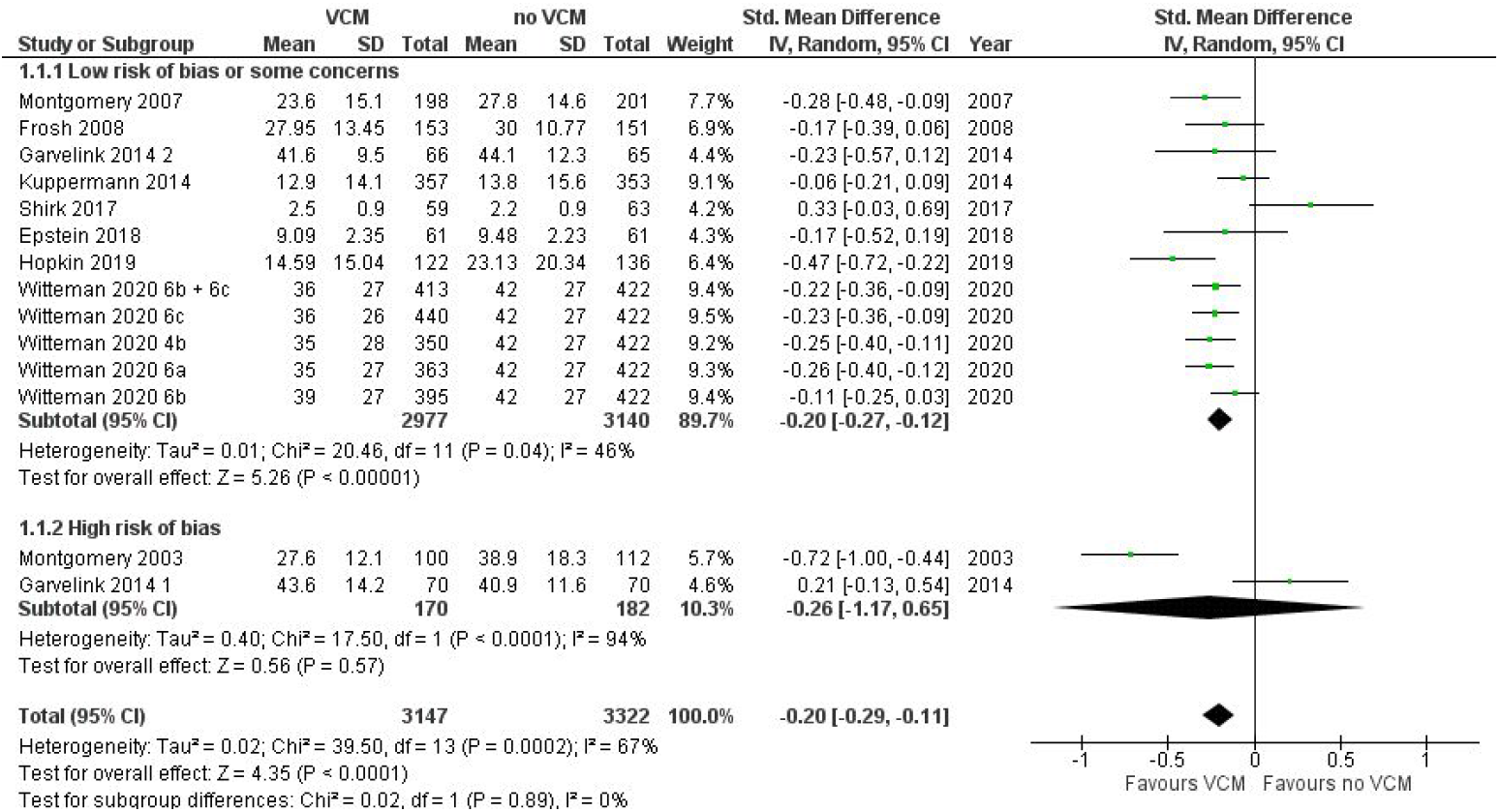
Decisional Conflict by Risk of Bias

#### Head-to-Head Evaluations of Values Clarification Methods

Feldman-Stewart et al. (2006) found no difference across all three groups (information only; values clarification method without a summary bar, i.e. rating scales; values clarification with a summary bar, i.e. multicriteria decision analysis) in terms of the attributes participants identified as important to their decisions nor in how difficult it was to make the decision. When trial participants were unblinded at the end of the study and shown all three options, all of them ranked the bars with the summary option (multicriteria decision analysis) as the most helpful.

Pignone et al. (2012) found that a discrete choice experiment produced somewhat different patterns of attribute importance compared to ranking and rating. Agreement between the most important attribute derived from the values clarification method and the most important attribute as reported by participants in the questionnaire was slightly higher in the ranking and rating arm than the discrete choice experiment arm. The authors found no difference between study arms in terms of values clarity, intent to be screened and unlabelled screening test preference.

Pignone et al. (2013) found that different values clarification methods produced differences in attribute importance and screening test preference. Participants who received the rating and ranking test were more likely to report the chance of dying from prostate cancer as the most important attribute compared to the balance sheet and discrete choice experiment groups. Those who received the balance sheet were more likely to prefer the unlabelled PSA-like test option compared to the two other groups.

Participants who received the discrete choice experiment were somewhat less likely to select reduction of mortality as the most important attribute, and were least likely to select the PSA-like option on the unlabelled preference question. There was no difference across groups in intent to be screened (labelled PSA test option) nor on values clarity.

Brenner et al. (2014) found that different values clarification methods produced different results in terms of individuals’ most important screening test attributes. Specifically, respondents who received the rating and ranking exercise, compared to a discrete choice experiment or a balance sheet (i.e., implicit values clarification method), were the most likely to choose risk reduction as the most important attribute. They found no differences in terms of test preferences, values clarity, nor intention to be screened.

Witteman et al. (2020) found that overall, methods using mathematical models (e.g., decision analysis, allocation of points) were more promising than other methods (e.g., pros and cons, rating scales) for encouraging values-congruent decisions. All methods encouraged lower decisional conflict when this was assessed.

#### Risk of Bias

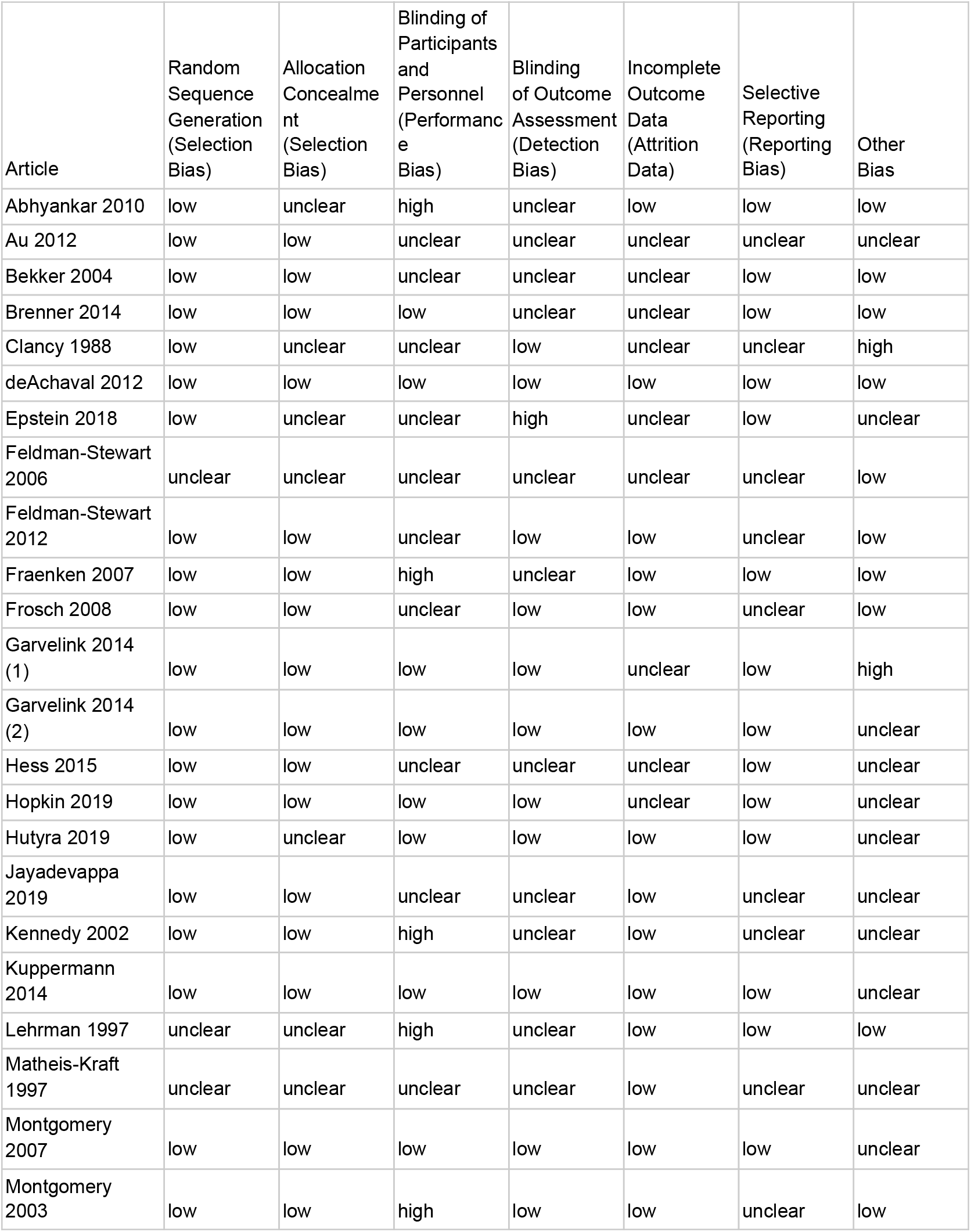
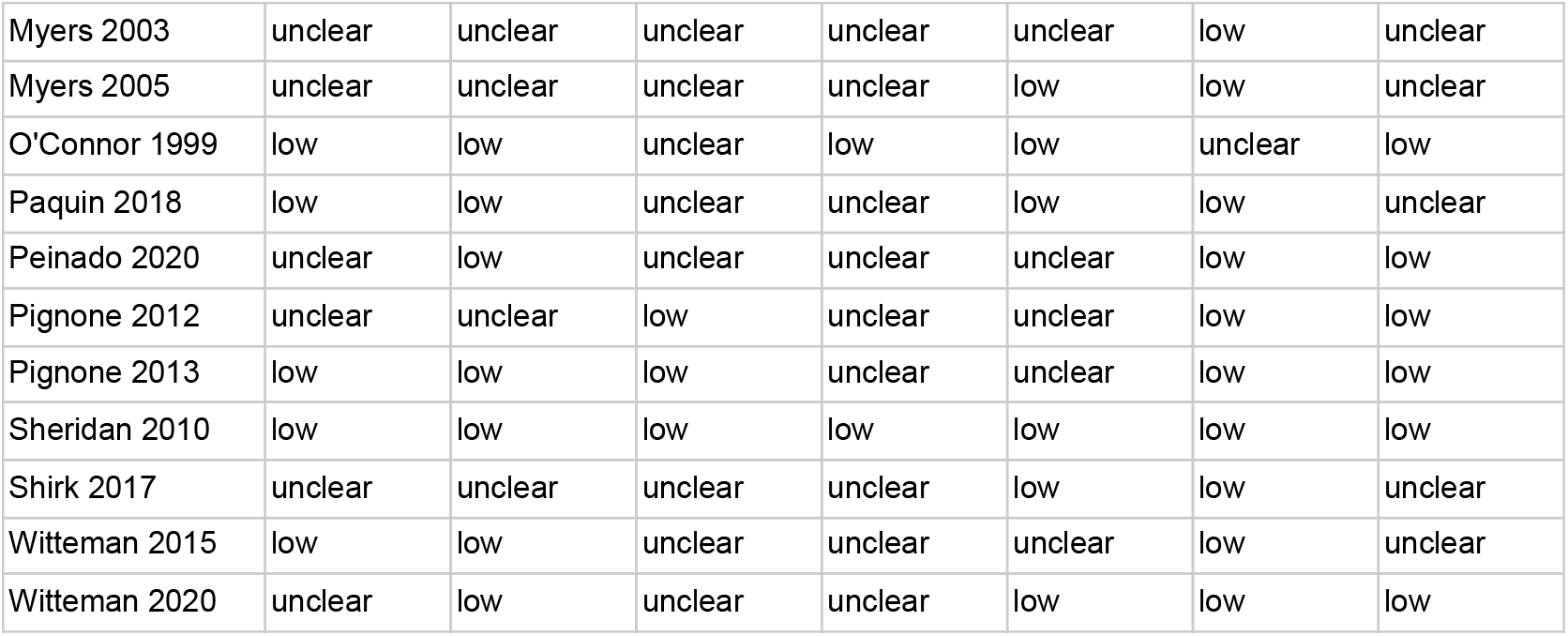

| Article | Random Sequence Generation (Selection Bias) | Allocation Concealment (Selection Bias) | Blinding of Participants and Personnel (Performance Bias) | Blinding of Outcome Assessment (Detection Bias) | Incomplete Outcome Data (Attrition Data) | Selective Reporting (Reporting Bias) | Other Bias |
| --- | --- | --- | --- | --- | --- | --- | --- |
| Abhyankar 2010 | low | unclear | high | unclear | low | low | low |
| Au 2012 | low | low | unclear | unclear | unclear | unclear | unclear |
| Bekker 2004 | low | low | unclear | unclear | unclear | low | low |
| Brenner 2014 | low | low | low | unclear | unclear | low | low |
| Clancy 1988 | low | unclear | unclear | low | unclear | unclear | high |
| deAchaval 2012 | low | low | low | low | low | low | low |
| Epstein 2018 | low | unclear | unclear | high | unclear | low | unclear |
| Feldman-Stewart 2006 | unclear | unclear | unclear | unclear | unclear | unclear | low |
| Feldman-Stewart 2012 | low | low | unclear | low | low | unclear | low |
| Fraenken 2007 | low | low | high | unclear | low | low | low |
| Frosch 2008 | low | low | unclear | low | low | unclear | low |
| Garvelink 2014 (1) | low | low | low | low | unclear | low | high |
| Garvelink 2014 (2) | low | low | low | low | low | low | unclear |
| Hess 2015 | low | low | unclear | unclear | unclear | low | unclear |
| Hopkin 2019 | low | low | low | low | unclear | low | unclear |
| Hutyra 2019 | low | unclear | low | low | low | low | unclear |
| Jayadevappa 2019 | low | low | unclear | unclear | low | unclear | unclear |
| Kennedy 2002 | low | low | high | unclear | low | unclear | unclear |
| Kuppermann 2014 | low | low | low | low | low | low | unclear |
| Lehrman 1997 | unclear | unclear | high | unclear | low | low | low |
| Matheis-Kraft 1997 | unclear | unclear | unclear | unclear | low | unclear | unclear |
| Montgomery 2007 | low | low | low | low | low | low | unclear |
| Montgomery 2003 | low | low | high | low | low | unclear | low |
| Myers 2003 | unclear | unclear | unclear | unclear | unclear | low | unclear |
| Myers 2005 | unclear | unclear | unclear | unclear | low | low | unclear |
| O'Connor 1999 | low | low | unclear | low | low | unclear | low |
| Paquin 2018 | low | low | unclear | unclear | low | low | unclear |
| Peinado 2020 | unclear | low | unclear | unclear | unclear | low | low |
| Pignone 2012 | unclear | unclear | low | unclear | unclear | low | low |
| Pignone 2013 | low | low | low | unclear | unclear | low | low |
| Sheridan 2010 | low | low | low | low | low | low | low |
| Shirk 2017 | unclear | unclear | unclear | unclear | low | low | unclear |
| Wittman 2015 | low | low | unclear | unclear | unclear | low | unclear |
| Wittman 2020 | unclear | low | unclear | unclear | low | low | low |

## Online Appendix 4: PRISMA Checklist

| Section/topic | # | Checklist item | Reported on page # |
| --- | --- | --- | --- |
| **TITLE** |  |  |  |
| Title | 1 | Identify the report as a systematic review, meta-analysis, or both. | Title page |
| **ABSTRACT** |  |  |  |
| Structured summary | 2 | Provide a structured summary including, as applicable: background; objectives; data sources; study eligibility criteria, participants, and interventions; study appraisal and synthesis methods; results; limitations; conclusions and implications of key findings; systematic review registration number. | 2 |
| **INTRODUCTION** |  |  |  |
| Rationale | 3 | Describe the rationale for the review in the context of what is already known. | 3-5 |
| Objectives | 4 | Provide an explicit statement of questions being addressed with reference to participants, interventions, comparisons, outcomes, and study design (PICOS). | 3-5 |
| **METHODS** |  |  |  |
| Protocol and registration | 5 | Indicate if a review protocol exists, if and where it can be accessed (e.g., Web address), and, if available, provide registration information including registration number. | na |
| Eligibility criteria | 6 | Specify study characteristics (e.g., PICOS, length of follow-up) and report characteristics (e.g., years considered, language, publication status) used as criteria for eligibility, giving rationale. | 6 |
| Information sources | 7 | Describe all information sources (e.g., databases with dates of coverage, contact with study authors to identify additional studies) in the search and date last searched. | 6-7 |
| Search | 8 | Present full electronic search strategy for at least one database, including any limits used, such that it could be repeated. | Appendix 1 |
| Study selection | 9 | State the process for selecting studies (i.e., screening, eligibility, included in systematic review, and, if applicable, included in the meta-analysis). | 7 |
| Data collection process | 10 | Describe method of data extraction from reports (e.g., piloted forms, independently, in duplicate) and any processes for obtaining and confirming data from investigators. | 7 |
| Data items | 11 | List and define all variables for which data were sought (e.g., PICOS, funding sources) and any assumptions and simplifications made. | 7 - 8 |
| Risk of bias in individual studies | 12 | Describe methods used for assessing risk of bias of individual studies (including specification of whether this was done at the study or outcome level), and how this information is to be used in any data synthesis. | 8 |
| Summary measures | 13 | State the principal summary measures (e.g., risk ratio, difference in means). | 8-9 |
| Synthesis of results | 14 | Describe the methods of handling data and combining results of studies, if done, including measures of consistency (e.g., $I^2$) for each meta-analysis. | 8-9 |
| Risk of bias across studies | 15 | Specify any assessment of risk of bias that may affect the cumulative evidence (e.g., publication bias, selective reporting within studies). | 8 |
| Additional analyses | 16 | Describe methods of additional analyses (e.g., sensitivity or subgroup analyses, meta-regression), if done, indicating which were pre-specified. | 8-9 |
| **RESULTS** |  |  |  |
| Study selection | 17 | Give numbers of studies screened, assessed for eligibility, and included in the review, with reasons for exclusions at each stage, ideally with a flow diagram. | 9 |
| Study characteristics | 18 | For each study, present characteristics for which data were extracted (e.g., study size, PICOS, follow-up period) and provide the citations. | 10-14 |
| Risk of bias within studies | 19 | Present data on risk of bias of each study and, if available, any outcome level assessment (see item 12). | 14 |
| Results of individual studies | 20 | For all outcomes considered (benefits or harms), present, for each study: (a) simple summary data for each intervention group (b) effect estimates and confidence intervals, ideally with a forest plot. | 14-17 |
| Synthesis of results | 21 | Present results of each meta-analysis done, including confidence intervals and measures of consistency. | 14-17 |
| Risk of bias across studies | 22 | Present results of any assessment of risk of bias across studies (see Item 15). | Appendix 3 |
| Additional analysis | 23 | Give results of additional analyses, if done (e.g., sensitivity or subgroup analyses, meta-regression [see Item 16]). | 14-17 |
| **DISCUSSION** |  |  |  |
| Summary of evidence | 24 | Summarize the main findings including the strength of evidence for each main outcome; consider their relevance to key groups (e.g., healthcare providers, users, and policy makers). | 17-18 |
| Limitations | 25 | Discuss limitations at study and outcome level (e.g., risk of bias), and at review-level (e.g., incomplete retrieval of identified research, reporting bias). | 18-19 |
| Conclusions | 26 | Provide a general interpretation of the results in the context of other evidence, and implications for future research. | 19 |
| **FUNDING** |  |  |  |
| Funding | 27 | Describe sources of funding for the systematic review and other support (e.g., supply of data); role of funders for the systematic review. | 20 |

*From:* Moher D, Liberati A, Tetzlaff J, Altman DG, The PRISMA Group (2009). Preferred Reporting Items for Systematic Reviews and Meta-Analyses: The PRISMA Statement. PLoS Med 6(7): e1000097. doi:10.1371/journal.pmed1000097

## Notes

### Competing Interest Statement

The authors have declared no competing interest.

### Author Declarations

N/A (systematic review)

